# Oncogenicity Variant Interpreter (OncoVI): oncogenicity guidelines implementation to support somatic variants interpretation in precision oncology

**DOI:** 10.1101/2024.10.10.24315072

**Authors:** Maria Giulia Carta, Lars Tögel, Annett Hölsken, Christoph Schubart, Heinrich Sticht, Robert Stöhr, Silvia Spoerl, Norbert Meidenbauer, Arndt Hartmann, Paolo Magni, Florian Haller, Fulvia Ferrazzi

## Abstract

**Background:** In precision oncology, the accurate and reproducible classification of variant oncogenicity is fundamental for therapy decision making. In 2022, a set of guidelines for the classification of oncogenicity of somatic variants in cancer were defined by the Clinical Genome Resource, the Cancer Genomics Consortium, and the Variant Interpretation for Cancer Consortium. However, to date an implementation that automates the evaluation of these criteria does not exist. Furthermore, for the majority of the criteria, the interpretation of the criterion-associated textual indication and the choice of publicly available resources to gather criterion-supporting information depends on the user. Thus, the risk of a variant being classified differently by independent laboratories is relevant, with implications for the management of patient care.

**Methods:** Here, we developed Oncogenicity Variant Interpreter (OncoVI), a fully-automated Python-based implementation of the oncogenicity guidelines. First, each criterion was interpreted and publicly available resources were identified to be utilised as reference. Then, criteria were implemented in OncoVI and the information reported by the associated resources were integrated. OncoVI is part of a broader Python-framework that, starting from the genomic position of the variant, automatically performs functional annotation, collects the available evidence from the reference resources, and provides a classification of oncogenicity.

**Results:** On the set of 93 somatic variants provided by the guidelines OncoVI achieved an overall accuracy of 80%, with a sensitivity of 88% in the classification of Oncogenic/Likely Oncogenic variants. On a real-world data set of 7,802 variants from 557 patients previously evaluated within the Molecular Tumour Board (MTB) Erlangen, an agreement of 79% was observed between OncoVI’s oncogenicity classification and the MTB pathogenicity assessment. In addition, the pathogenicity classification of 135 variants of the MTB data set was re-assessed by expert biologists adhering solely to the oncogenicity guidelines. This re-evaluation confirmed the validity of OncoVI’s interpretation of the resources chosen as reference, but it also underlined the ability of experts in solving conflicting evidence.

**Conclusions:** Taken together, OncoVI provides an effective implementation of the oncogenicity guidelines, thus facilitating their adoption and supporting the reproducible and harmonised oncogenicity classification of somatic variants between institutions.

## Background

Precision oncology aims at identifying personalised cancer treatments on the basis of patient-specific tumour mutations (1). To establish precision oncology as routine practice there are two prerequisites: mutation identification via next-generation-sequencing (NGS) of cancer cell DNA, and precise, reproducible classification of the oncogenicity of somatic variants and clinical interpretation in the context of the disease. Nowadays, the comprehensive analysis of tumour profiles is a widely consolidated practice in the clinical setting, facilitated by the accessibility of NGS technologies and their cost reduction (2, 3). Tumour profiling has so far been mainly addressed through targeted panel sequencing, but whole exome and whole cancer genome sequencing have recently emerged as advanced diagnostic tools in personalised medicine (4–8).

While the establishment of the pathogenicity of a variant aims at determining the impact of the variant on protein function, the classification of oncogenicity of a somatic variant aims at clarifying its role in cancer initiation and progression. To this aim, guidelines for the correct classification of variant pathogenicity and actionability have been proposed (9, 10). However, the harmonisation across laboratories remains challenging, with still open questions and sensitive consequences of clinical relevance (11–13). The process of oncogenicity classification can be articulated in three main steps: functional annotation, collection of biomedical evidence, and final interpretation. Functional annotation predicts the effect of the nucleotide change on protein level. Well-established, widely used tools capable of returning predictions for a single variant in a fraction of a second currently exist, such as Variant Effect Predictor (VEP) (14), Annovar (15), and SnpEff (16). The collection of biological and clinical evidence requires the identification and integration of different resources, like for example population data, biomedical literature, and in-silico predictions. For this purpose, tools such as VarSome (17) and cBio Cancer Genomics Portal (cBioPortal) (18) exist. These tools integrate and display different data types in an intuitive way, but leave the responsibility for the final interpretation to the user. The main challenges in the interpretation of somatic variants are the choice of the appropriate resources to use, out of all those available, and the integration of the different information they provide. To support the laborious task of interpreting the evidence collected for genomic variants in cancer, many projects have focused on the development of databases containing curated information, like the Clinical Interpretation of Variants in Cancer (CIViC), the Oncology Knowledge Base (OncoKB), the Database of Curated Mutations (DoCM), and the Cancer Genome Interpreter (CGI) (19–22). Their contribution is precious as they are able to provide a classification of pathogenicity, oncogenicity or clinical significance. However, the information they contain is predominantly limited to well-known and characterised genes. Moreover, scientific indications are reported for only a minority of variants and they are heterogeneous in content (23). Recently, machine-learning methods, such as AlphaMissense (24), have been proposed to effectively support the prediction of the impact of unknown variants on protein function and their potential role in cancer. Although very valuable in the research field, their results have only a supporting role in the clinical context, because they cannot be used as the sole evidence to draw scientific conclusions without experimental validation. Thus, the choice, integration, and interpretation of biomedical resources are still open challenges for the correct oncogenicity classification of somatic variants in cancer.

In order to increase the agreement in variant interpretation between different institutions as well as between different assessors, a harmonisation of the oncogenicity classification is needed. To this aim, in 2022 the Clinical Genome Resource (ClinGen), the Cancer Genomics Consortium (CGC), and the Variant Interpretation for Cancer Consortium (VICC) published standards for the classification of oncogenicity of somatic variants in cancer (25). Based on a Standard Operating Procedure (SOP) of 17 criteria, provided as textual indications, somatic single nucleotide variants and small insertions/deletions are classified into one of five classes, i.e., Oncogenic, Likely Oncogenic, Variant of Uncertain Significance (VUS), Likely Benign, and Benign. For some criteria careful interpretation by the user is essential, such as for criteria that refer to well-established in vitro or in vivo functional studies supportive of an oncogenic or benign effect of the variant. Furthermore, the authors provide recommendations for the use of resources for a few but not all criteria.

Here, we take a further step towards the harmonisation of the interpretation of somatic variants by presenting Oncogenicity Variant Interpreter (OncoVI), a tool providing a fully automated evaluation of the oncogenicity guidelines. OncoVI is incorporated in a broader Python-based framework that, taking as input the genomic coordinates of the variants, (i) performs functional annotation, (ii) collects the available clinical evidence according to the interrogated resources, and (iii) evaluates each criterion providing a final classification of oncogenicity. OncoVI thus facilitates the adoption of the oncogenicity guidelines, allowing the standardisation of the oncogenicity classification of somatic variants across institutions.

## Material and Methods

### Standard Operating Procedure data set

The Standard Operating Procedure (SOP) data set contains 93 variants and was obtained as the manually curated union of supplementary tables 1 and 3 provided by the authors of the oncogenicity guidelines (**Additional File 1**) (25). The 93 variants are located in 10 well-known genes: i.e., six oncogenes (OGs), three tumour supressor genes (TSGs), and one gene with a context-dependent role. The 70 missense variants in supplementary table 3 were provided with the following information for both genome reference builds 37 and 38: Gene, DNA change (c.), AA change 3 letter code, AA change 1 letter code, DNA change (g.). In addition, chromosome, genomic position, strand, reference base and alternate base were provided for both genome reference builds 37 and 38. For the remaining 23 variants only Gene, DNA change, AA change 3 letter code, and AA change 1 letter code were available in supplementary table 1. Thus, chromosome, genomic position, strand, reference base and alternate base were derived manually from the DNA change. Furthermore, for all variants the triggered criteria, the variant-specific scores, and the final oncogenicity classification were available in the corresponding supplementary tables, published by the authors of the guidelines, and manually collected (**Additional File 1**).

### Molecular Tumour Board data set

The Molecular Tumour Board (MTB) data set is composed of 7,802 unique somatic variants obtained from real-world routine diagnostic data of 557 patients with various tumour diagnoses (**Supplementary Figure 3A**), enrolled in the setting of the MTB of the CCC Erlangen-EMN (Germany) from February 2022 to February 2024. Formalin-fixed paraffin embedded patient tissue was sequenced on a NextSeq500 using the targeted sequencing TruSight Oncology 500 assay (TSO500, Illumina, Inc., San Diego, CA, USA), which investigates 523 cancer-related genes. Sequencing raw data (bcl files) were processed and analysed utilising the TruSight Oncology 500 v2.2 Local App provided as Docker container on a remote server based on Ubuntu’s 20.04.6 long-term support (LTS) operating system. The pipeline includes demultiplexing and fastq generation, DNA and RNA alignment to the human reference genome (UCSC University of California Santa Cruz, hg19), calling of single and multiple nucleotide variants, copy number variants, fusions, and splice variants. Genomic positions of the variants detected in each patient were provided as output in a Variant Call Format (VCF) file called MergedSmallVariants.genome.vcf. The classification of pathogenicity for each variant in one of five classes (i.e., Pathogenic, Likely Pathogenic, Variant of Uncertain Significance, Likely Benign, Benign) of the variants had been manually curated by three expert cancer biologists.

### MTB validation data set

In addition to the SOP data set and the MTB data set, a validation data set of 135 somatic variants was created from the MTB data set to validate the implementation of the oncogenicity’s criteria in OncoVI. The set of 135 variants was created by merging selected (n=15) and randomly chosen variants (n=120) with a potentially critical discrepancy between the MTB pathogenicity and OncoVI’s oncogenicity assessment (see Results for a detailed description) (**Additional File 5**). The same three expert cancer biologists who manually curated the classification of pathogenicity of the MTB data set were engaged to classify the subset of 135 somatic variants according to the oncogenicity criteria. To this aim, the validation data set was randomly divided into three separate sets and a predefined set of resources, corresponding to those implemented in OncoVI, was provided to the experts as reference for each criterion.

### Variant annotation via VEP-based in-house pipeline

Functional annotation of the variants in the SOP data set (**Additional File 2**) and of the variants in the MTB data set (**Additional File 3**) was performed relying on a custom Python-based pipeline (Python version 3.8.8), running in a dedicated conda environment on a remote server based on Ubuntu’s 20.04.6 long-term support (LTS) operating system. The pipeline takes the genomic positions of the variants as input. For the SOP data set, the coordinates in the hg38 reference genome build for the 70 missense variants (provided in supplementary table 3 of the original publication) were utilised, whereas for the remaining 23 variants genomic coordinates were manually derived from the DNA change (provided in supplementary table 1 of the original publication) were used. For the variants in the MTB data set, hg19 genomic coordinates were obtained by uplifting of the original VCFs (i.e., MergedSmallVariants.genome.vcf) to the genome reference build hg38 via the *LiftOverVcf* function of the picard package (version 3.0.0).

As first step, the Python-based annotation pipeline applies hard filters, i.e., only genomic coordinates with an alternate allele different from the reference allele are selected and only variants with FILTER equal to PASS or Blacklist are retained using the function filter_vcf of the VcfFilterPy package (https://github.com/superDross/VcfFilterPy). In a second step, variants that survived the hard filters are functionally annotated with the VEP from Ensembl (VEP version 111.0, January 2024) utilising GRCh38 and Reference Sequence (RefSeq) Database (26) as preferred cache. Two VEP plugins were employed to retrieve scores from predictive algorithms during the annotation step, namely dbNSFP (database of non-synonymous functional predictions, version 4.5a) (27) and spliceAI (28). The third step consists in post-processing the VCF annotated by VEP. Next, the filter_vep tool was used to remove from the VCF the variants without Consequence and/or a global gnomAD (29) population frequency ≥0.025. In addition, in the post-processing step only variants for which VEP had annotated a coding consequence were retained. Furthermore, for variants for which multiple transcripts were annotated, the canonical one according to the MANE project (30) was favoured. As final step, the pipeline interrogates several knowledgebases to retrieve existing biological and clinical information of the somatic variants. For the purposes of this study, only the pathogenicity classification and review level of the ClinVar database (31) were exploited via the ClinVar API. As final output, the pipeline provides a list of all the annotated variants that underwent the abovementioned steps in the form of tabular data (csv and Microsoft Excel).

### Implementation of the oncogenicity’s criteria in OncoVI

All the criteria for oncogenic and benign effect as well as the point-based system for the classification of oncogenicity of somatic variants from the SOP proposed by Horak et al. were implemented in a Python-based framework (version 3.8.8), except for the “Oncogenic Supporting-2” (OP2) criterion. First, for each criterion, provided as textual indication, its biological meaning had to be interpreted. For example, according to our interpretation, the criterion “Oncogenic Moderate-1” (OM1, 2 points, “*Located in a critical and well-established part of a functional domain*”) for evidence of oncogenic effect addressed variants located in a functional domain of the protein. As by definition a protein’s domain folds and functions independently from the rest of the polypeptide chain, we hypothesised that variants affecting protein domains are more likely to disrupt the protein function. Then, publicly available resources and data types to be utilised for each criterion were either adopted from those suggested in the SOP or had to be identified among publicly available databases, when a suggestion by the SOP was not present. For example, for OM1 the identified resource was UniProt and human protein domains were employed. Each criterion was translated into an IF-ELSE statement, on which OncoVI relies to determine if the information, gathered from the identified resource, supports the evidence, and assigns the corresponding pre-defined points accordingly. For example, OM1 was implemented in OncoVI as follows: first OncoVI verifies the existence in UniProt of a protein encoded by the gene on which the variant under evaluation is located, and IF the triplets of nucleotides affected by the variant belongs to a functional domain, THEN OM1 is triggered, otherwise not. The sum of the pre-defined points associated with the triggered criteria, hereafter referred to as the variant-specific score, is used as a measure of oncogenicity for the evaluated variant. Full details of the resources used and levels of evidence considered to trigger each criterion are described in the Supplementary materials. **Additional File 1** and **Additional File 4** contain OncoVI classification of the SOP and MTB data sets, respectively.

### Data visualisation and statistical analyses

All statistical analyses and figures were generated within R version 4.1.2/ Bioconductor version 3.13 environment (32). Spearman correlations were computed with the function *cor.test()* of the stats package. Contingency tables were generated using the R package caret version 6.0.94. All plots were created with the R packages ggplot2 version 3.5.1 and ggalt version 0.4.0. OncoPrints were produced with the R package ComplexHeatmap version 2.13.1. Alluvial plots was created using the R package ggalluvial version 0.12.5.

## Results

The SOP for oncogenicity classification of somatic variants by Horak et al. provides 12 and five precise and consistent criteria for the point-based evaluation of the oncogenic effect and benign effect, respectively. For each somatic variant, all criteria are evaluated and a variant-specific score is calculated. On the basis of this score the variant is classified into one of five possible classes, i.e., Oncogenic, Likely Oncogenic, Variant of Uncertain Significance (VUS), Likely Benign, and Benign. Our OncoVI tool was designed in order to be an easy-to-use software offering an automated evaluation of the oncogenicity criteria (**Figure 1**). First, each criterion needed to be interpreted; then publicly available resources were chosen as reference for each criterion, either relying on the SOP suggestions or identifying them from publicly available databases when no suggestion by the SOP was provided. As in the SOP, the oncogenicity classification was based on the calculated variant-specific score.

**Figure 1.**
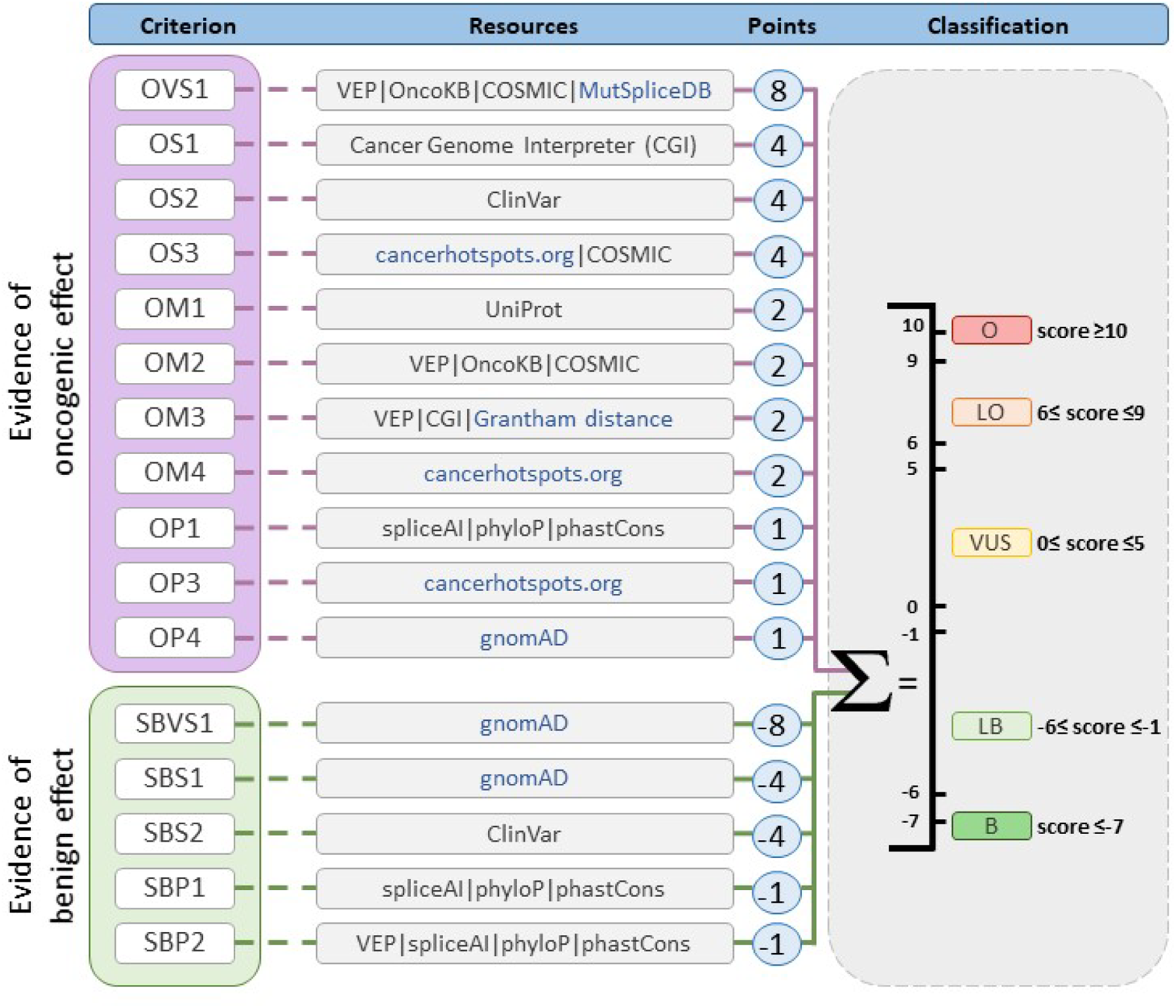
Workflow of OncoVI. The figure shows the implemented criteria in OncoVI (11 and five criteria for evidence of oncogenic and benign effect respectively), the public resources utilised to assess each criterion, the points associated with each criterion and the classification of oncogenicity into one of five classes on the basis of the variant-specific score, obtained as the sum of the points associated to the criteria triggered by OncoVI for the variant: scores≥10:Oncogenic (O), 6≤scores≤9:Likely Oncogenic (LO), 0≤scores≤5:Variant of uncertain significance (VUS), −6≤score≤-1:Likely Benign (LB), scores≤-7: Benign (B). Blue: resources suggested by the authors of the guidelines, black: resources identified by the authors of this study.

### Performance of OncoVI on the Standard Operating Procedure data set

To assess the validity of the resources utilised for each criterion and of our automated interpretation of the oncogenicity’s criteria, OncoVI was tested on the SOP data set provided by the oncogenicity guideline authors (25). The data set included 93 somatic variants, for which the variant-associated triggered criteria, the variant-specific scores, and the oncogenicity classifications were available. Variants in the SOP data set were located in ten different well-known cancer-related genes, i.e., six oncogenes (OGs) (i.e., *KRAS*, *BRAF*, *PIK3CA*, *IDH1*, *TERT, FLT3*), three TSGs (i.e., *PTEN*, *TP53*, and *RB1*), and *EZH2* whose role in cancer is context-dependent. Variants of the SOP data set were distributed across the five oncogenicity classes as follows: Oncogenic: 14, Likely Oncogenic: 29, VUS: 38, Likely Benign: 6, and Benign: 6 (**Additional File 1**). Variant annotation via our in-house VEP-based pipeline showed that the most prevalent type of variants were missense (n=71), truncating (n=9) and variants affecting the upstream gene region (n=5) (**Additional File 2**).

To evaluate OncoVI’s performance on the SOP data set, we grouped Likely Oncogenic and Oncogenic classes into the class Oncogenic/Likely Oncogenic (O/LO) and Likely Benign and Benign classes into another class Benign/Likely Benign (B/LB). When considering the SOP’s oncogenicity classification as ground truth based on three oncogenicity classes (i.e., O/LO, VUS and B/LB), the overall accuracy was 80% (n=74/93 correctly classified variants), with a sensitivity of 88% for the O/LO class (n=38/43) and a sensitivity of 83% for the B/LB class (n=10/12) (**Figure 2A**). OncoVI classified five O/LO variants as VUS: three occurring in the *EZH2* gene, the other two in the promoter region of gene *TERT*. Furthermore, we observed two B/LB variants classified as VUS, one missense variant in the *TP53* gene and the other in the promoter region of *TERT*. Taken together, OncoVI showed very good agreement with the SOP in terms of variant oncogenicity classification.

**Figure 2.**
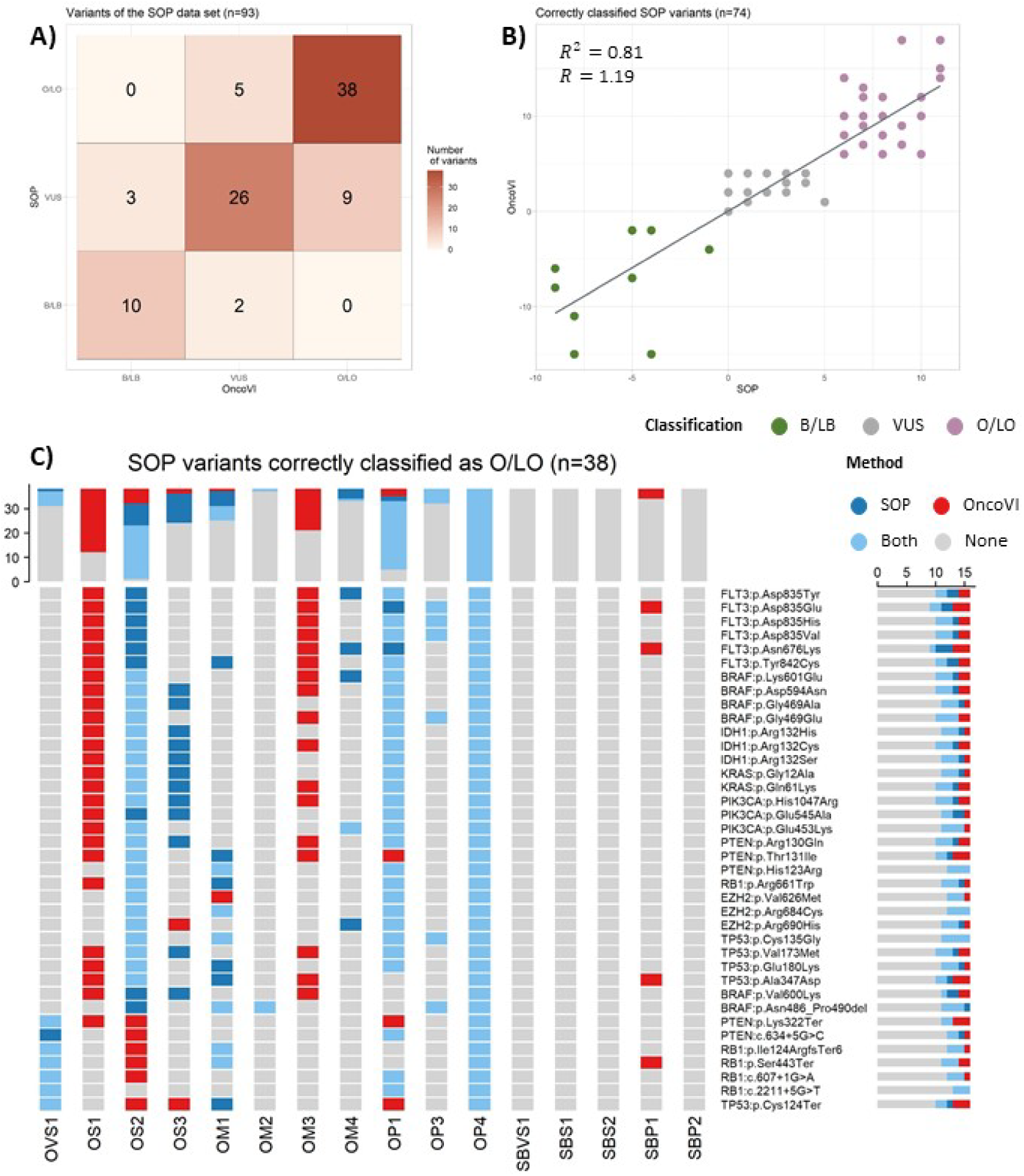
Results of OncoVI on the Standard Operating Procedure (SOP) data set. **A)** Confusion matrix of the agreement between the SOP and OncoVI in classifying the somatic variants of the SOP data set. B/LB: Benign/Likely Benign. VUS: Variant of uncertain significance. O/LO: Oncogenic/Likely Oncogenic. **B)** Scatterplot between the points assigned by the SOP and the points assigned by OncoVI for the 74 correctly classified variants R^2^: coefficient of determination: R: regression coefficient **C)** Oncoprint of the 38 variants of the SOP data set correctly classified as O/LO. Each row corresponds to one variant, each column corresponds to an assessed criterion The colour of the cell indicates whether a criterion was triggered by the SOP only (dark blue), OncoVI only (red), both (11 ght blue), or none of the two (grey). The top barplot shows the sum of cells of each colour calculated across all variants, the barplot on the right shows the sum across all criteria. OVS1: Oncogenic Very Strong-1 (8 points), OS1: Oncogenic Strong-1 (4 points), OS2: Oncogenic Strong-2 (4 points), OS3: Oncogenic Strong-3 (4 points), OM1: Oncogenic Moderate-1 (2 points), OM2: Oncogenic Moderate-2 (2 points), OM3: Oncogenic Moderate-3 (2 points), OM4: Oncogenic Moderate-4 (2 points), OP1: Oncogenic Supportiıg-1 (1 point), OP3: Oncogenic Supporting-3 (1 point), OP4: Oncogenic Supporting-4 (1 point), SBVS1: Somatic Benign Very Strong-1 (−8 points), SBS1: Somatic Benign Strong-1 (−4 points), SBS2: Somatic Benign Strong-2 (−4 points), SBP1: Somatic Benign Supporting-1 (−1 point), SBP2: Somatic Benign Supporting-2 (−1 point).

Next, the agreement between the SOP and OncoVI in terms of variant-specific scores was evaluated. When considering the 74 correctly classified variants in the SOP data set, a strong positive correlation (0.87, p-value < 2.2e-16) between the points assigned by the SOP and by OncoVI was observed (**Figure 2B**). Furthermore, when the scores assigned by the SOP and OncoVI differed, OncoVI assigned in most cases higher scores than the SOP in correctly classified O/LO (**Supplementary Figure 1A**) and in correctly classified VUS variants (**Supplementary Figure 2C**), and lower scores in correctly classified B/LB variants (**Supplementary Figure 2A**). Overall OncoVI assigned higher points than the SOP in correctly classified variants, which aligns with our aim to have a high sensitivity for the O/LO class.

To further compare OncoVI and the SOP, the most frequently criteria triggered by OncoVI in the 74 correctly classified variants were investigated. The three most frequently triggered criteria in the set of 38 variants correctly classified as O/LO were “Oncogenic Supporting-4” (OP4, 1 point), “Oncogenic Supporting-1” (OP1, 1 point), and “Oncogenic Strong-2” (OS2, 4 points) (**Supplementary Figure 1B**). Furthermore, “Oncogenic Very Strong-1” (OVS1, 8 points, “*Null variants in a bona fide TSG*”) was triggered in six cases. Four variants triggered “Somatic Benign Supporting-1” (SBP1, −1 point) for evidence of benign effect, because these variants were predicted as not potentially harmful by the computational algorithms used as a reference. In the ten variants correctly classified as B/LB, SBS2 (Strong, −4 points) was the top triggered criterion. This criterion is triggered if a variant is reported in ClinVar as Benign/Likely Benign with a submission method supported by functional studies (see Supplementary Material for details). “Somatic Benign Strong-1” (SBS1, −4 points) and “Somatic Benign Very Strong-1” (SBVS1, −8 points), which apply when the variant is found in the population database gnomAD in at least one of the five general continental populations at a minor allele frequency (MAF) >1% and >5%, respectively, were triggered by seven and four variants, respectively (**Supplementary Figure 2B**). In the 26 variants correctly classified as VUS, only criteria of category Supporting [OP4: n=26, OP1: n=20, “Oncogenic Supporting-3” (OP3): n=3] and of category Moderate (OM1: n=15) for evidence of oncogenic effect were triggered (**Supplementary Figure 2D**). Overall, in the correctly classified O/LO variants, OncoVI triggered criteria for oncogenic effect of category Very Strong (8 points) and Strong (4 points), which are associated with the highest points and thus contribute the most to the variant-specific score. Analogously, in correctly classifying B/LB variants OncoVI chose criteria for benign effect of category Very Strong (−8 points) and Strong (−4 points), which are the most influential criteria in lowering the variant-specific score for correct benign classification. Taken together, the assessment of the triggered criteria supported the appropriateness of the resources chosen as reference, and showed that the classification of oncogenic and benign variants mainly depends on triggering of the respective strong criteria.

To identify potential reasons for the higher scores assigned by OncoVI, we investigated the criteria triggered by the SOP and OncoVI focusing individually on each of the 38 variants correctly classified as O/LO (**Figure 2C**).

The strongest agreement between the two methods in the triggered criteria was observed for OP4 (n=38/38 variants), OP1 (n=28/38 variants), OS2 (n=22/38 variants), and OVS1 (n=6/38 variants). Instead, OS1 (Strong, 4 points, “*Same amino acid change as a previously established oncogenic variant (using this standard) regardless of nucleotide change*”) was exclusively triggered by OncoVI. This is likely due to the fact that, according to our interpretation, OS1 required a set of previously established oncogenic variants, based on the oncogenicity criteria themselves. Since this set is not available when applying the guidelines for the first time, OncoVI’s implementation relied on an external data set of validated oncogenic mutations (i.e., the Catalog of Validated Oncogenic Mutations of the GCI database). Analogously, also OM3 (Moderate, 2 points, “*Missense variant at an amino acid residue where a different missense variant determined to be oncogenic (using this standard) has been documented*”) was exclusively applied by OncoVI. Furthermore, the oncogenicity guidelines provided an exception to the applicability of OS3 (Strong, 4 points), which is the activation of OS1 for the same variant under consideration. Thus, as a consequence of the activation of OS1 by OncoVI alone, OS3 was triggered almost exclusively by the SOP (n=12/38 variants). Taken together, for the variants correctly classified as O/LO, OncoVI showed good agreement with the SOP in activating the criteria, with the only exception of two criteria where the differences were most likely due to the choice of utilising an external reference data set of oncogenic variants. Overall, the results of the comparison between OncoVI and the SOP confirmed the validity of the automated implementation offered by OncoVI.

### Performance of OncoVI on the Molecular Tumour Board data set

To evaluate the performance of OncoVI in a real-world setting, we utilised a data set of 7,802 unique somatic variants previously classified in the context of our MTB, Erlangen by three expert cancer biologists. The 7,802 variants (hereafter referred to as “MTB data set”) had been detected in a cohort of 557 patients with different tumour entities (**Supplementary Figure 3A**) in 489 different genes. Differently from the SOP data set, here no clear indication about the role of the genes as either OG or TSG or context-dependent was available, thus making the MTB data set not only larger but also more heterogeneous. Another key difference is that variants of the MTB data set had been assessed for pathogenicity, defined as the deleteriousness or activating effect of the variant on the protein’s function, without specifically considering the neoplastic context as in the assessment of oncogenicity. Indeed, the 7,802 variants had been assigned to one of five classes of pathogenicity (i.e., Pathogenic, Likely Pathogenic, VUS, Likely Benign, and Benign), (**Supplementary Figure 3B**, **Additional File 4**), without utilising the oncogenicity guidelines outlined in the SOP. To perform a comparison between the MTB assessment and OncoVI, the variants belonging to Pathogenic and Likely Pathogenic classes were grouped into one class hereafter referred to as the “Pathogenic/Likely Pathogenic” (P/LP) class. In addition, the variants classified as Benign or Likely Benign were grouped together into another class called “Benign/Likely Benign” (B/LB).

Variants in the MTB data set were annotated with our in-house VEP-based pipeline and interpreted with OncoVI. Functional annotation revealed that the three most frequent type of variants were missense (n=6,363), frameshift (n=540), and stop gained (n=467) (**Additional File 3**). The performance of OncoVI was evaluated by assessing the agreement of its oncogenicity classification into O/LO, VUS, and B/LB with the expert biologists’ pathogenicity classification into P/LP, VUS, and B/LB. The observed agreement was 78% (n=6,058/7,802 variants), in line with the accuracy observed for the SOP data set (80%, n=74/93 variants). In particular, VUS and B/LB classes had an agreement of respectively 97% (n=3,956/4,092 variants) and 43% (n=961/2,261 variants), while the agreement between OncoVI’s O/LO and MTB P/LP was 79% (n=1,141/1,449 variants) (**Figure 3A**). Two variants (the missense variant *HNF1A*:p.Pro379His and the donor region variant *HNRNPK*:c.516+3A>T) classified as P/LP by the MTB assessment were classified by OncoVI as B/LB. Both variants triggered SBS2 (Strong, −4 points, “*Well-established in vivo or in vitro functional studies showing no oncogenic effect*”), because they were reported in ClinVar with a Benign and Likely Benign effect, respectively at the time of submissions (December 2023). However, the expert pathogenicity assessment was carried out at an earlier date, thus with insufficient data leading to a misinterpretation. Conversely, 13 variants classified as B/LB according to the MTB assessment were classified by OncoVI as O/LO. Ten out of these 13 misclassified variants triggered OVS1 (Very Strong, 8 points, “*Null variant in a bona fide tumor suppressor gene*”), while the remaining three activated a criterion of category Strong (4 points) and two criteria of category Supporting (1 point) for oncogenic effect, which explained the oncogenicity classification as Likely Oncogenic by OncoVI. Investigation into the factors underlying the classification of these 13 variants revealed that they were classified by the MTB as B/LB more likely either because they (i) were located in the terminal part of the of the protein or (ii) had a conflicting interpretation of pathogenicity in ClinVar, or (iii) were located on a splicing site.

**Figure 3.**
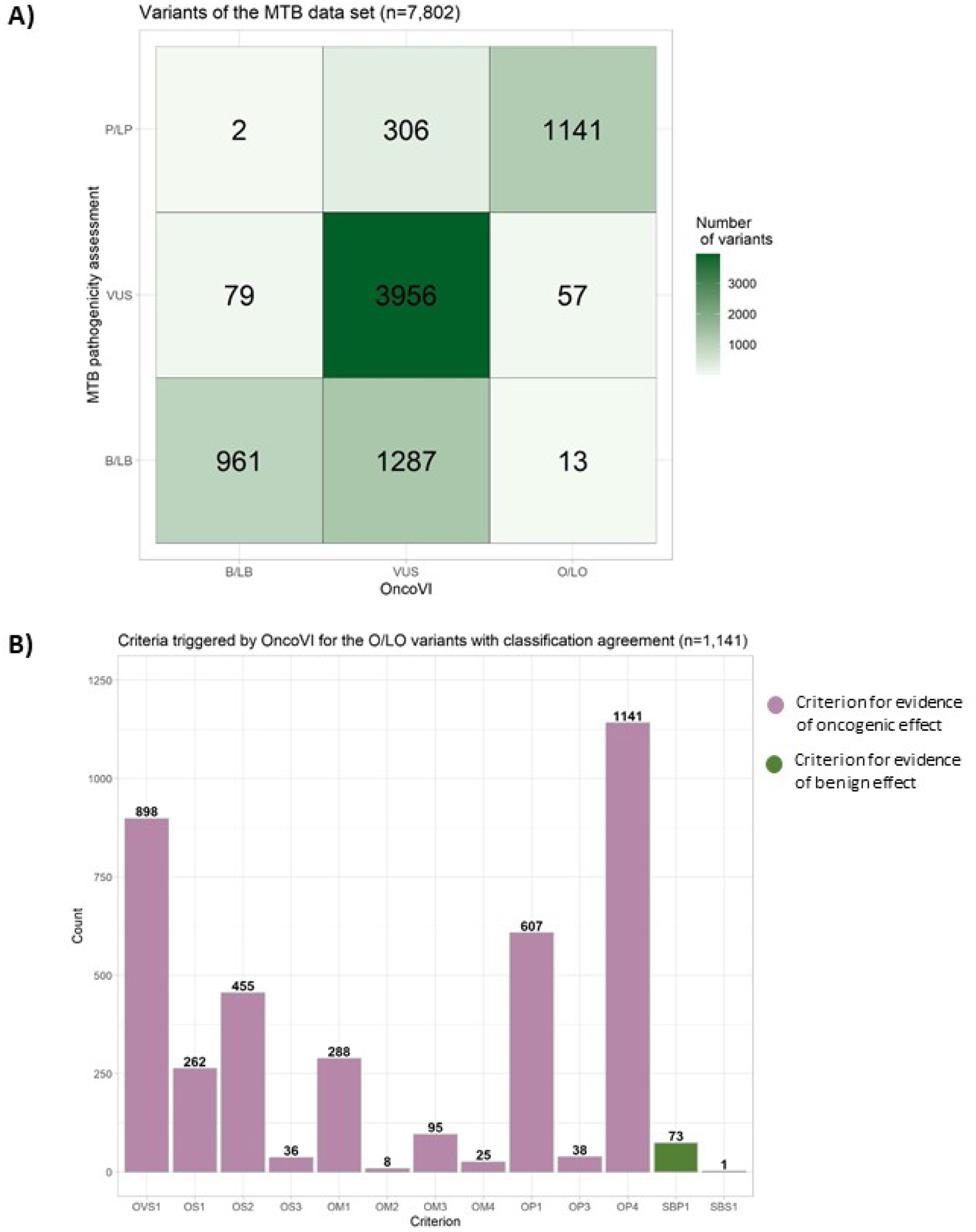
Results of OncoVI on the Molecular Tumour Board (MTB) data set. **A)** Confusion matrix of the agreement between the MTB pathogenicity assessment and OncoVI’s oncogenicity in classifying the 7,802 variants of the MTB data set. B/LB: Benign/Likely Benign. VUS: Variant of uncertain significance. O/LO: Oncogenic/Likely Oncogenic P/LP: Pathogenic/Likely Pathogenic. **B)** Barplot of the criteria triggered by OncoVI in the 1.141 variants with classification agreement between MTB pathogenicity and OncoVI’s oncogenicity assessment. Criteria are sorted according to decreasing corresponding points. OVS1: Oncogenic Very Strong-1 (8 points), OS1: Oncogenic Strong-1 (4 points), OS2: Oncogenic Strong-2 (4 points), OS3: Oncogenic Strong-3 (4 points), OM1: Oncogenic Moderate-1 (2 points), OM2: Oncogenic Moderate-2 (2 points). OM3: Oncogenic Moderate-3 (2 points). OM4: Oncogenic Moderate-4 (2 points). OP1: Oncogenic Supporting-1 (1 point), OP3: Oncogenic Supporting-3 (1 point), OP4: Oncogenic Supporting-4 (1 point), SBP1: Somatic Benign Supporting-1 (−1 point), SBS1: Somatic Benign Strong-1 (−4 points)

As the most critical issue for the use of OncoVI to support oncogenicity classification are false negative O/LO variants, the 306 P/LP variants classified as VUS by OncoVI were examined in further detail. The majority of these variants (64%, n=196/306) had a consequence calculated by our in-house VEP-based pipeline categorised as truncating (i.e., frameshift, stop gained) or affecting the splicing site of the protein. This would explain why these variants were classified as P/LP in the MTB, while a possible reason why OncoVI did not classify them as O/LO is that the application of OVS1 was not triggered because they were not localised in a bona fide TSG.

Furthermore, as for the SOP data set, the most frequently criteria triggered by OncoVI were investigated. In the 1,141 variants with agreement between MTB pathogenicity and OncoVI’s oncogenicity classification the top three most frequently triggered criteria were: OP4, OVS1, and OP1 for evidence of oncogenic effect (**Figure 3B**).

As for the SOP data set, OP4 (Supporting, 1 point, “*Absent from controls (or at an extremely low frequency) in gnomAD*”) was the top triggered criterion. OVS1 (Very Strong, 8 points, “*Null variants in a bona fide TSG*”) was triggered in 79% (n=898/1,141) of the variants, thus revealing a higher percentage of TSGs in the real-world MTB data set than in the SOP data set (16%, n=6/38). The benign criterion SBP1 (Supporting, −1 point) was triggered 73 times, because the computational algorithms used as reference for SBP1 predicted a benign effect of these variants. In the 961 B/LB variants with agreement between MTB and OncoVI’s oncogenicity classification, SBS2 (Strong, −2 points, “*Well-established in vitro or in vivo functional studies showing no oncogenic effects*”) was the most frequently triggered criterion by OncoVI (n=821/961) (**Supplementary Figure 4A**). This further confirmed the appropriateness of ClinVar as reference for SBS2 (Strong, −4 points). SBP1 (n=549), SBS1 (n=496), and SBVS1 (n=149) for evidence of benign effect were also frequently triggered. In the 3,956 VUS variants with agreement between MTB pathogenicity and OncoVI’s oncogenicity classifications, OP4, OP1 and OM1 for evidence of oncogenic effect (OP4: n=3,956, OP1: n=2,988, OM1: n=1,760) were the first three most frequently criteria triggered by OncoVI. SBP1 was triggered for 790 variants because the computational algorithms used as reference reported a benign effect (**Supplementary Figure 4B**). Taken together, despite the discrepancies due to the difference between pathogenicity and oncogenicity assessments, results showed good overall agreement between OncoVI and MTB variant classification. In addition, OncoVI correctly triggered criteria of strong evidence for oncogenic effect and of strong evidence for benign effect in O/LO and B/LB variants respectively, with agreement in MTB pathogenicity classification.

### Comparative analysis with expert re-assessment of the MTB data set

As the expert cancer biologists had not employed the oncogenicity’s guidelines in the assessment of the variants within the framework of the MTB, a comparison with OncoVI was only possible in terms of pathogenicity classification. Therefore, to more extensively assess the performance of OncoVI in classifying the oncogenicity of somatic variants in a real-world scenario, the expert cancer biologists re-assessed a subset of these somatic variants solely adhering to the oncogenicity guidelines and employing the same resources used by OncoVI. Yet, the experts were blinded to the IF-ELSE translation of the criteria implemented in OncoVI, and no suggestion on the data type to use was given. The subset of re-assessed variants, hereafter referred to as “validation data set”, was chosen on the basis of the results of the comparison between OncoVI and the MTB assessment (**Figure 3A**) in order to (i) identify potential reasons for the observed discrepancies in the classification performances and (ii) assess whether the implementation of the criteria in OncoVI is consistent with the human’s interpretation of the resources. Specifically, the 135 variants of the validation data set included the two variants classified within the MTB as P/LP and misclassified by OncoVI as B/LB, the 13 variants classified within the MTB as B/LB and misclassified by OncoVI as O/LO, and randomly chosen variants both from those with classification agreement between MTB pathogenicity and OncoVI’s oncogenicity and with discrepancies in the classification (**Supplementary Figure 5A**, **Additional File 5**). Expert classification based on the oncogenicity guidelines had an agreement of 34% (n=46/135 variants) with their previous pathogenicity assessment in the MTB (**Figure 4A**, **Supplementary Figure 5B**). Specifically, four variants were confirmed as B/LB (14%, n=4/28) and 26 variants previously classified within the MTB as P/LP were confirmed O/LO (34%, n=26/77), while all the remaining B/LB and P/LP variants were re-assessed as VUS.

**Figure 4.**
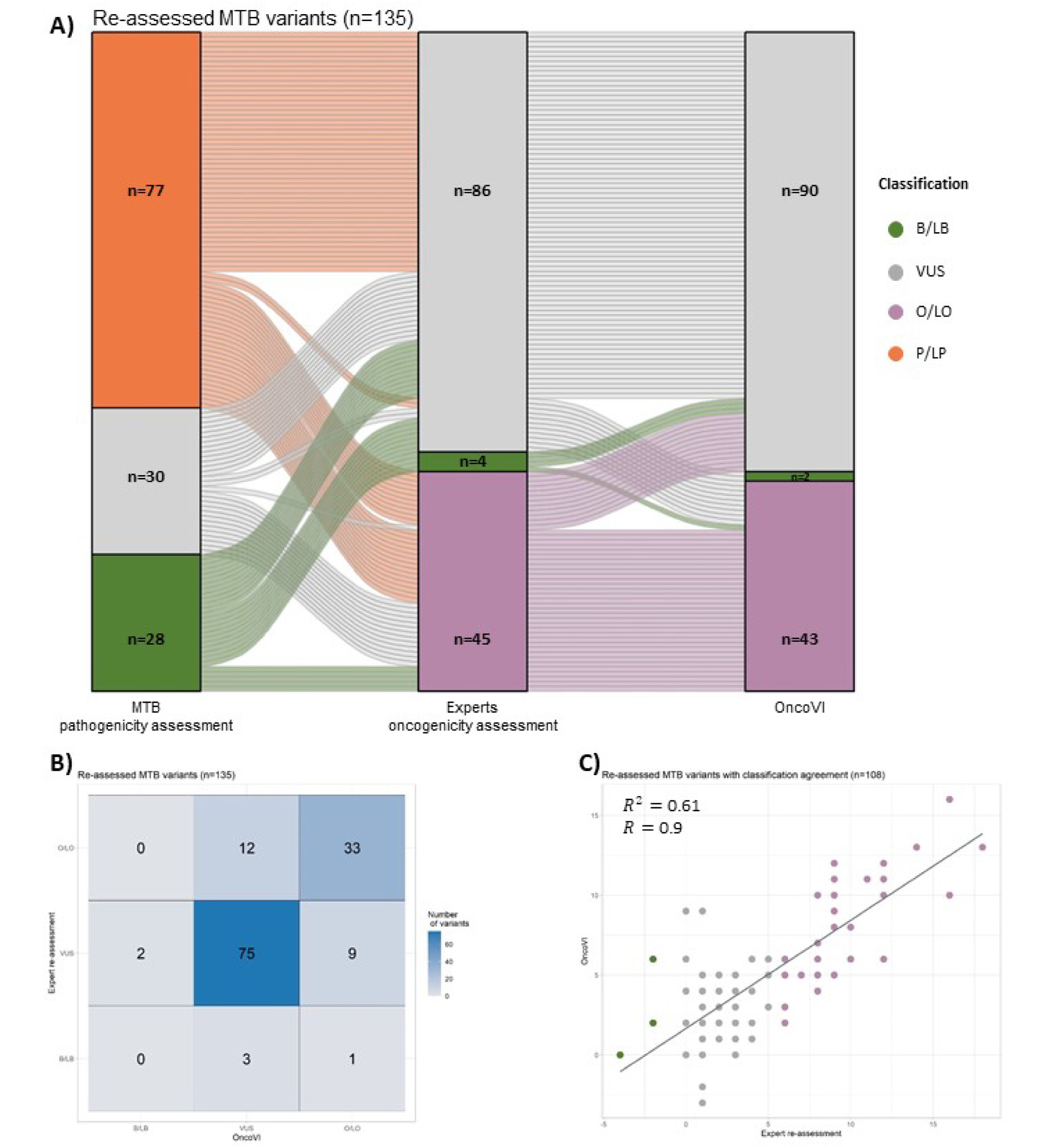
Results of OncoVI on the subset of MTB variants re-assessed by experts based on oncogenicity. **A)** Alluvial plot of the 135 variants of the validation data set re-assessed by experts based on oncogenicity. Horizontally-distributed columns (axes) represent variants classified by the pathogenicity assessment within the Molecular Tumour Board (MTB) (left), by the re-assessment of the experts based on the oncogenicity guidelines (middle), and by OncoVI (right) Alluvial flows between axes show whether variants maintain or change their classification. B/LB: Benign/Likely Benign. VUS: Variant of uncertain significance. O/LO: Oncogenic/Likely Oncogenic. P/LP: Pathogenic/Likely Pathogenic. **B)** Confusion matrix of the agreement between OncoVI and the expert re-assessment, based on the oncogenicity guidelines, in classifying the somatic variants of the validation data set **C)** Scatterplot between the points assigned by the experts and the points assigned by OncoVI for the 108 variants with agreement between expert and OncoVI’s oncogenicity classification R2: coefficient of determination; R: regression coefficient

The agreement of OncoVI with the pathogenicity classification performed within the MTB was 22% (n=30/135 variants) (**Supplementary Figure 5A**) and was 80% (n=108/135 variants) with experts’ re-assessment (**Figure 4B**). No variants assessed by experts as O/LO were classified as B/LB by OncoVI, yet one variant (*PALB2*:p.Leu939Trp) re-assessed by experts as B/LB was classified by OncoVI as O/LO. This is due to the presence of this variant in the CGI database (OS1, 4 points), to a prediction of deleteriousness by computational algorithms (OP1, 1 point), and to the presence at extremely low frequency in gnomAD (OP4, 1 point). (**Additional File 5**). Taken together, while more agreement is expected when experts specifically follow the guidelines, the strong observed increase in agreement between OncoVI and experts’ assessment strengthened the hypothesis that most discrepancies emerged between OncoVI and MTB classification were due to assessing pathogenicity and not oncogenicity.

In addition to the comparison in terms of oncogenicity’s classification, the concordance in terms of variant-specific scores assigned by OncoVI and the expert re-assessment was also evaluated as done for the SOP data set. When considering the 108 correctly classified variants, a positive correlation of 0.7 (p-value < 2.2e-16) between the scores was observed (**Figure 4C**).

For 13 out of 33 O/LO variants with agreement between expert and OncoVI’s oncogenicity classification, OncoVI and the expert re-assessment attributed the same number of points, while for the remaining 20 variants OncoVI assigned in most cases lower scores (**Supplementary Figure 6A**, **Additional File 5**). When focusing on the 75 VUS variants with agreement between expert and OncoVI’s oncogenicity classification, 32 of them showed the same scores in both methods and in 28 of the remaining 43 variants OncoVI assigned higher scores than experts (**Supplementary Figure 7A**, **Additional File 5**). Two of the four variants re-assessed by experts as B/LB were classified by OncoVI as VUS since no criteria were triggered (**Additional File 5**). In summary, these results showed that the concordance between OncoVI and expert re-assessment was also reflected by the assignment of comparable scores to the variants.

Next, the most frequently triggered criteria were evaluated. In the 33 O/LO variants with agreement between expert and OncoVI’s oncogenicity classification, the two criteria most frequently triggered were OP4 (n=33/33) and OVS1 (n=25/33) for evidence of oncogenic effect (**Supplementary Figure 6B**). In the VUS variants with agreement between expert and OncoVI’s oncogenicity classification, the top three most triggered criteria were of category Supporting and Moderate (OP4: n=75/75; OP1: n=34/75, OM1: n=34/75) (**Supplementary Figure 7B**). Criteria for evidence of benign effect, i.e., SBP1 and SBS2, were triggered in ten and one cases, respectively. Overall, as observed in the SOP data set, the results on the validation data set confirmed OncoVI’s ability to activate strong criteria for oncogenic effect that have thus a strong influence on correct oncogenicity classification of O/LO variants.

To more closely investigate how the criteria implemented in OncoVI emulate the human expert interpretation of the resources chosen as reference, we individually examined all the criteria triggered by the experts and OncoVI in the 108 variants with oncogenicity classification agreement (**Supplementary Figure 8**). Criteria where experts and OncoVI showed the strongest agreement were: OP4 (triggered by both in 107 variants), OVS1 (n=26 variants), OS1 (n=7) and OM2 (n=8). Of note, 9 variants where both methods triggered OVS1 (Very Strong, 8 points, “*Null variants in a bona fide TSG*”) had been classified as VUS by the MTB assessment, but re-classified as O/LO by experts and OncoVI. Although truncating, seven of them were located in the terminal exons of the protein, which is the reason why experts classified these variants within the MTB as VUS. OS2 (Strong, 4 points, “*Well-established in vivo or in vitro functional studies showing oncogenic effect of the variant*”) and SBS2 (Strong, −4 points, “*Well-established in vivo or in vitro functional studies showing no oncogenic effect of the variant”*) which used ClinVar as resource were among the criteria more frequently applied by experts alone (respectively 12 and four times). These variants reported either “Conflicting interpretation” in ClinVar, or a classification of pathogenicity or benign effect but without submission methods supported by functional studies (see Supplementary Material for details). This observation underlines the ability of experts to solve conflicting evidence compared to OncoVI’s automated implementation, and might be another reason for the observed discrepancies between MTB and OncoVI classifications. Criteria with strongest disagreement between OncoVI and experts were OM1, OP1, and SBP1. In OM1 (Moderate, 2 points, “*Variant located in a well-established part of a functional domain*”), UniProt is used as reference. Here, OncoVI and experts agreed on n=41/108 variants, while in 13 and 16 cases the criterion was exclusively triggered by experts or OncoVI only, suggesting UniProt as resource may not be suitable for automated interpretation. OP1 (Supporting, 1 point) and SBP1 (Supporting, −1 point) are criteria considering predictions of oncogenic versus benign effect from computational algorithms (i.e., phyloP100way_vertebrate_rankscore, phastCons100way_vertebrate_rankscore, spliceAI). OP1 was triggered by both methods in n=25/108 variants, by OncoVI alone in n=37/108 variants and by experts alone in n=14/108 variants. Conversely, SBP1 was triggered by both in n=6/108 variants, applied exclusively by OncoVI none times and by experts eight times. This marked difference could be due to experts’ and OncoVI’s use of different online resources to retrieve the predictive scores as well as different interpretation of these predictions (e.g. different chosen thresholds for the deleterious/tolerated prediction). Collectively, these results showed that OncoVI and experts strongly agreed in triggering criteria not requiring interpretation of the resources (e.g., OP4, OVS1, OS1, OS3, OM2, OP3). In contrast, the discordance between the two methods in criteria such as OS2, OM1, OP1, SBS2 and SBP1 was likely due to the expert interpretation of the resources.

## Discussion

The precise and reproducible oncogenicity classification of somatic variants still represents an open issue in precision oncology. In 2022, a Standard Operating Procedure (SOP) consisting of 17 criteria was defined to standardise this classification across institutions. However, the criteria were provided as textual indications, and only for a subset of them a resource was suggested to be used as reference. Here, we developed Oncogenicity Variant Interpreter (OncoVI), an easy-to-use Python-based tool to automate the oncogenicity guideline evaluation. OncoVI has the potential to broaden the adoption of the SOP, further contributing to the standardisation of the oncogenicity classification. In OncoVI each criterion was interpreted, associated with a publicly available resource, and then translated to an IF-ELSE statement. The variants-specific score, i.e., the sum of the points associated with the triggered criteria, allowed the classification of oncogenicity.

Testing OncoVI on the SOP data set of variants revealed high agreement with the SOP’s oncogenicity classification. Furthermore, focusing on concordantly classified variants, the variant-specific scores assigned by OncoVI strongly correlated with those assigned by the manual adoption of the SOP, though OncoVI’s scores were in general higher. This, however, aligns with our intention to minimise the chance to miss real oncogenic variants even at the potential cost of more false positive oncogenic calls to be revised manually by experts. The ability of OncoVI in triggering criteria of category Strong for oncogenic or benign effects, in variants correctly classified as O/LO and B/LB respectively, confirmed the correct interpretation of the implemented criteria as well as the appropriateness of the chosen resources. In addition, also at the level of the individual triggered criteria, good consistency between OncoVI and the SOP was observed, with few exceptions linked to the choice to utilise an external database of previously classified oncogenic variants as reference for two criteria in OncoVI. The triggering of these criteria by OncoVI alone might also be a reason for the higher variant-specific scores assigned by OncoVI.

When testing OncoVI on a real-world Molecular Tumour Board (MTB) data set, comprising 7,802 variants, the observed agreement between MTB pathogenicity assessment and OncoVI was about 80%. This result is in line with the accuracy observed for the SOP data set, despite the fact that the MTB data set contains a more heterogeneous set of genes and a higher proportion of missense variants, whose role in cancer development is more challenging to unravel (33). Notably, the classification in the context of the MTB was based on pathogenicity and thus differed from the oncogenicity criteria employed by OncoVI and the SOP. However, the results still showed high agreement between MTB pathogenicity and OncoVI’s oncogenicity assessments. As for the SOP data set, also for the MTB data set we observed a higher agreement between OncoVI’s O/LO variants and MTB P/LP variants, than in the B/LB class. The oncogenicity classification by OncoVI of the two P/LP MTB variants as B/LB was most likely due to changing evidence in ClinVar, used as resource. However, during expert re-assessment based on the oncogenicity guidelines these two variants were re-evaluated as VUS, based on solely applying criterion OP4 (Supporting, 1 point, “*Absent from controls (or at an extremely low frequency) in gnomAD*”) in the oncogenicity assessment of these variants. This highlights that variant assessment may change over time according to revised content of the respective resource and further stresses the importance of an automated implementation to ensure reproducibility in the variant interpretation process. These aspects are of crucial importance contributing to the well-known issue of variant re-classification (34). On the other hand, investigating the factors underlying the classification by OncoVI of B/LB MTB variants as O/LO revealed a limitation of OncoVI’s automated implementation. For example, we observed the difficulty of OncoVI in solving conflicting classifications of pathogenicity in ClinVar as well as the correct classification of either variants affecting the splicing site or truncating mutations in the terminal exons of the gene, which most likely do not affect protein functionality, such as stop codons in the last exon as they are unlikely to activated nonsense mediated mRNA decay (35). Thus, special attention should be paid during the evaluation of this type of variants. In addition, a likely key reason for the observed discrepancies between OncoVI and MTB classifications, in particular the high proportion of P/LP variants classified by OncoVI as VUS, was the intrinsic difference between the pathogenicity assessment by the MTB and the oncogenicity evaluation performed by the criteria implemented in OncoVI. Indeed, pathogenicity evaluates the deleteriousness of the variant to the protein function without considering the neoplastic context. Therefore, the truncating or splice-affecting nature of these types of variants led experts to classify them as pathogenic in the MTB, which is different from classifying them as oncogenic. As in the SOP data set, OncoVI triggered strong oncogenic and benign criteria in the MTB variants with classification agreement. Yet, some differences also emerged. For example, the observed higher proportion of variants in the MTB data set triggering OVS1 may be explained by the higher prevalence of mutated tumour suppressor genes (TSGs) than oncogenes (OGs) or genes whose role is unclear in real-world data sets. In addition, the less frequent activation of criteria that rely on curated databases, such as OS2 and OS1, could be due to database limitations in reporting variant information for only well-characterised genes. This suggests that curated databases may not be of sufficient support for rare variant interpretation. To mitigate this issue, integration of text mining of medical literature as source of evidence for the guidelines will be considered in the future. In contrast, the comparable frequency of SBS2 activation in the two data sets may be related to the higher presence of variants classified as B/LB than P/LP in ClinVar.

The re-assessment of a subset of MTB variants by expert assessors strengthened the hypothesis that several discrepancies emerged in the MTB data set were attributable to the difference in pathogenicity and oncogenicity evaluations. In addition, the disagreement between OncoVI and experts in a minority of re-assessed variants further highlighted the limitations of an automated implementation in dealing with conflicting interpretation in ClinVar. To mitigate this issue, flagging variants in OncoVI that have a conflicting interpretation could be considered, so that they can be manually revised.

Collectively, our results show that OncoVI is a valuable tool to automate the interpretation of oncogenicity guidelines in somatic variants. In addition, OncoVI is provided as open-source software in the widely used programming language Python to allow its customisation and integration into pre-existing pipelines. However, this study also highlights that OncoVI is a tool that cannot fully substitute human expertise and therefore should be used as a tool to support molecular biologists and physicians in the oncogenicity classification of somatic variants. Furthermore, OncoVI is intended for research purposes only and its use outside of research context is under the responsibility of the user, who should also comply with licences of the resources utilised.

## Conclusions

Collectively, our results show that OncoVI effectively automates and streamlines the evaluation of the oncogenicity guidelines, thus supporting the standardisation and reproducibility of oncogenicity classification in precision oncology.

## Data Availability

All data produced in the present work are contained in the manuscript

## List of abbreviations

B: Benign
B/LB: Benign/Likely Benign
cBioPortal: cBio Cancer Genomics Portal
CGC: Cancer Genomics Consortium
CGI: Cancer Genome Interpreter
CIViC: Clinical Interpretation of Variants in Cancer
ClinGen: Clinical Genome Resource
COSMIC: Catalogue of Somatic Mutations in Cancer
dbNSFP: database of non-synonymous functional predictions
DoCM: Database of Curated Mutations
gnomAD: Genome Aggregation Database
Indel: insertion-deletion
LB: Likely Benign
LO: Likely Oncogenic
LTS: Long-Term Support
MAF: minor allele frequency
MTB: Molecular Tumour Board
NGS: next-generation-sequencing
O: Oncogenic
OG: oncogene
O/LO: Oncogenic/Likely Oncogenic
OM1: Oncogenic Moderate-1
OM3: Oncogenic Moderate-3
OP1: Oncogenic Supporting-1
OP2: Oncogenic Supporting-2
OP3: Oncogenic Supporting-3
OP4: Oncogenic Supporting-4
OncoKB: Oncology Knowledge Base
OncoVI: Oncogenicity Variant Interpreter
OS1: Oncogenic Strong-1
OS2: Oncogenic Strong-2
OS3: Oncogenic Strong-3
OVS1: Oncogenic Very Strong-1
P/LP: Pathogenic/Likely Pathogenic
RefSeq: Reference Sequence Database
SBP1: Somatic Benign Supporting-1
SBP2: Somatic Benign Supporting-2
SBS2: Somatic Benign Strong-2
SBVS1: Somatic Benign Very Strong-1
SNV: Single Nucleotide Variant
SOP: Standard Operating Procedure
TSG: tumour supressor gene
TSO500: TruSight Oncology 500
UCSC: University of California Santa Cruz
UniProt: Universal Protein Knowledgebase
VCF: Variant Call Format
VEP: Variant Effect Predictor
VICC: Variant Interpretation for Cancer Consortium
VUS: Variant of Uncertain Significance

## Declarations

### Ethics approval and consent to participate

The study was conducted in accordance with the Declaration of Helsinki and approved by the Ethics Committee of the Friedrich-Alexander University Erlangen-Nuremberg (100_17 B from 7 April 2017, addendum from 27 July 2021).

### Consent for publication

Not applicable.

### Availability of data and materials

All data generated or analysed during this study are included in this published article and its supplementary information files.

### Competing interests

ArHa obtains honoraria for lectures or consulting/advisory boards for Abbvie, Agilent, AstraZeneca, Biocartis, BMS, Boehringer Ingelheim, Cepheid, Diaceutics, Gilead, Illumina, Ipsen, Janssen, Lilly, Merck, MSD, Nanostring, Novartis, Pfizer, Qiagen, QUIP GmbH, Roche, Sanofi, 3DHistech and other research support from AstraZeneca, Biocartis, Cepheid, Gilead, Illumina, Janssen, Nanostring, Novartis, Owkin, Qiagen, QUIP GmbH, Roche, Sanofi. FH obtains honoraria for lectures or consulting/advisory boards for AstraZeneca, BMS, Boehringer Ingelheim, Novartis and receives research support from Illumina GmbH. The remaining authors have no financial or non-financial competing interests to declare.

### Funding

Supported by Deutsche Forschungsgemeinschaft (German Research Foundation) grant TRR 305 [projects Z01 (FF) and Z02 (ArHa)].

### Authors’ contributions

MGC and FF designed the study and wrote the manuscript. MGC implemented OncoVI, FF supervised OncoVI’s implementation. LT, AnHo, and CS interpreted variants of the MTB and validation data sets. HS, RS, SS, NM, and ArHA provided resources. MGC, LT, AnHo, CS, PM, FH, and FF interpreted the data. All authors read and approved the final manuscript.

## Acknowledgments

We express gratitude to patients, clinicians, technicians and institutions who contributed to the Molecular Tumour Board of the CCC Erlangen-EMN (Germany).

## Supplementary Material

### Criteria for evidence of oncogenicity of somatic variants

#### Oncogenic Very Strong-1 (OVS1) criterion

OVS1 (8 points) description is “*Null variants (nonsense, frameshift, canonical ±1 or 3 splice sites, initiation codon, single-exon or multiexon deletion) in a bona fide tumour suppressor genes*”. Null variants were defined relying on the calculated consequences by the Variant Effect Predictor (VEP) of Ensembl. “*nonsense”* corresponded to “stop_gained”, “stop_lost”, “*frameshift”* corresponded to “frameshift_variant”. *“canonical ±1 or 3 splice sites”* corresponded to either “splice_donor_variant” or “splice_acceptor_variant” or any consequence containing “splice” and affecting the 4 bases flanking the canonical sites. “*initiation codon”* corresponded to “start_lost”. “*single-exon or multiexon deletion*” were not implemented, given that VEP does not provide any consequence representing these variants. The set of TSGs was created as the union of two resources: the Cancer Gene Census (CGC) of the Catalogue of Somatic Mutations in Cancer (COSMIC) and OncoKB. From the CGC, genes with “Tier” equal to “1” and “Role in Cancer” equal to “TSG” were used (Census_allWed_May_1_17_27_56_2024.csv, v99, GRCh38, Accessed 1 May 2024) From OncoKB, genes in the Cancer Gene List (OncoKB-CGL) with “Is Tumor Suppressor Gene” equal to “Yes” was utilised (“cancerGeneList.tsv”, Accessed 29 April 2024).

For variants not triggering OVS1 MutSpliceDB (36) is consulted (“MutSpliceDB_BRP_2024-02-06.csv”, Accessed 06 February 2024).

#### Oncogenic Strong-1 (OS1) criterion

OS1 (4 points) description is “*Same amino acid change as a previously established oncogenic variant (using this standard) regardless of nucleotide change*”. A publicly available database of validated oncogenic variants was used as substitute for “*previously established oncogenic variant*”. Namely, the Catalog of Validated Oncogenic Mutations of the Cancer Gene Interpreter (GCI) database (Accessed 01 February 2024) was used. All variants regardless the context were considered.

#### Oncogenic Strong-2 (OS2) criterion

OS2 (4 points) description is “*Well-established in vitro or in vivo functional studies, supportive of an oncogenic effect of the variant*”. ClinVar API was exploted as reference for OS1. Variants with a clinical significance “Pathogenic” or “Likely pathogenic” and a submission method with the presence of functional studies were considered (i.e., “Practice guideline”, “Reviewed by expert panel”, “Criteria provided, multiple submitters, no conflicts”, “Criteria provided, single submitter”). Additionally, to trigger OS2, a curated list of clinical significances that are not matched by the abovementioned strings was used. To this aim, ClinVar database was downloaded (Accessed 28 February 2024) and 12 additional clinical significances were selected.

#### Oncogenic Strong-3 (OS3) criterion

OS3 (4 points) description is “*Located in one of the hotspots in* cancerhotspots.org *with at least 50 samples with a somatic variant at the same amino acid position, and the same amino acid change count in* cancerhotspots.org *in at least 10 samples*”. cancerhotspots.org was used (Accessed 26 September 2023). In case of cancerhotspots.org not reported variants at the residue in which was located the variant under evaluation, COSMIC Census Genes Mutations (Accessed 09 April 2024, v99) and COSMIC Cancer Mutation Census were considered (Accessed 07 May 2024, v99).

#### Oncogenic Moderate-1 (OM1) criterion

OM1 (2 points) description is “*Located in a critical and well-established part of a functional domain*”. For OM1, the bed file containing the human domain annotations from the UniProt database was downloaded (UP000005640_9606_domain.bed, release 2024_02, Accessed 16 April 2024). “UniProtKB_AC” (accession ids) were extracted and converted into HUGO gene symbols, via the webservice provided by UniProt, to be used as identifiers in the implementation.

#### Oncogenic Moderate-2 (OM2) criterion

OM2 (points) description is “*Protein length changes as a result of in-frame deletions/insertions in a known oncogene or tumor suppressor gene or stop-loss variants in a known tumor suppressor gene*”. For “*in-frame deletions/insertions”* VEP annotated consequences “inframe_deletion” and “inframe_insertion” were considered. To create TSGs and OGs groups OncoKB-CGL and COSMIC CGC genes, regardless of the Tier, were used.

#### Oncogenic Moderate-3 (OM3) criterion

OM3 (2 points) is “*Missense variant at an amino acid residue where different missense variant determined to be oncogenic (using this standard) has been documented. Amino acid difference should be greater or at least approximately the same as for missense change determined to be oncogenic*”. For missense variants VEP annotated consequence “missense_variant” was considered. The Catalog of Validated Oncogenic Mutations of GCI database was used as substitute for “*previously established oncogenic variant*”. To measure the difference between reference and alternate amino acids we used the Grantham’s distance (37).

#### Oncogenic Moderate-4 (OM4) criterion

OM4 (2 points) is “*Located in one of the hotspots in* cancerhotspots.org *with < 50 samples with a somatic variant at the same amino acid position, and the same amino acid change count in* cancerhotspots.org *is at least 10*”. As for OS3 criterion, cancerhotspots.org was used.

#### Oncogenic Supporting-1 (OP1) criterion

OP1 (1 point) description is “*All used lines of computational evidence support an oncogenic effect of a variant*”. Predictive scores from phyloP100way_vertebrate_rankscore (phyloP) (38) and phastCons100way_vertebrate_rankscore (phastCons) (39) were retrieved based on the VEP plugin dbNSFP (database of Non Synonymous Functional Prediction, version 4.5a). Additionally, scores from spliceAI were retrieved via the VEP plugin spliceAI. Variants with a phyloP or phastCons score ≥0.5, or a spliceAI score equal to “PASS” were considered to trigger the criterion.

#### Oncogenic Supporting-3 (OP3) criterion

OP3 (1 point) description is “*Located in one of the hotspots in* cancerhotspots.org *and the particular amino acid change count in* cancerhotspots.org *is below 10*”. As for OS3 and OM4 cancerhotspots.org was used.

#### Oncogenic Supporting-4 (OP4) criterion

OP4 (1 point) description is “*Absent from controls (or at an extremely low frequency) in gnomAD*”. gnomAD exome (gnomADe=“r2.1.1”) and genome frequency data are populated via VEP (gnomADg=“v3.1.2”). Variants absent in gnomAD exome (gnomADe) or genome (gnomADg) frequency data or with a gnomADe or gnomADg frequency value ≤0.01 trigger OP4.

#### Criteria for evidence of benign effect of somatic variants Somatic Benign Very Strong-1 (SBVS1) criterion

SBVS1 (−8 points) description is “*Minor allele frequency is >5% in gnomAD in any 5 general continental populations: African, East Asian, European (non-Finnish), Latino, and South Asian*”. The criterion is triggered by variants with a gnomAD frequency in at least one of the 5 populations ≥0.05.

#### Somatic Benign Strong-1 (SBS1) criterion

SBS1 (−4 points) description is “*Minor allele frequency is >1% in gnomAD in any 5 general continental populations: African, East Asian, European (non-Finnish), Latino, and South Asian*”. The criterion is triggered by variants with a gnomAD frequency in at least one of the 5 populations ≥0.01.

#### Somatic Benign Strong-2 (SBS2) criterion

SBS2 (−4 points) description is “*Well-established in vitro or in vivo functional studies show no oncogenic effects*”. Variants with a clinical significance “Benign” or “Likely benign” and a review status assuming a submission method reporting the presence of functional studies trigger SBS2.

#### Somatic Benign Supporting-1 (SBP1) criterion

SBP1 (−1 point) description is “*All used lines of computational evidence suggest no effect of a variant*”. phyloP100way_vertebrate_rankscore and phastCons100way_vertebrate_rankscore scores <0.5 or spliceAI score equal to “FAIL” trigger SBP1.

#### Somatic Benign Supporting-2 (SBP2) criterion

SBP2 (−1 point) description is “*A synonymous (silent) variant for which splicing prediction algorithms predict no effect on the splice consensus sequence nor the creation of a new splice site and the nucleotide is not highly conserved*”. For synonymous variants VEP calculated consequence equal to “synonymous_variant” were considered. phyloP100way_vertebrate_rankscore and phastCons100way_vertebrate_rankscore scores <0.5 or spliceAI score equal to “FAIL” trigger SBP2.

## Supplementary Figures

**Supplementary Figure 1.**
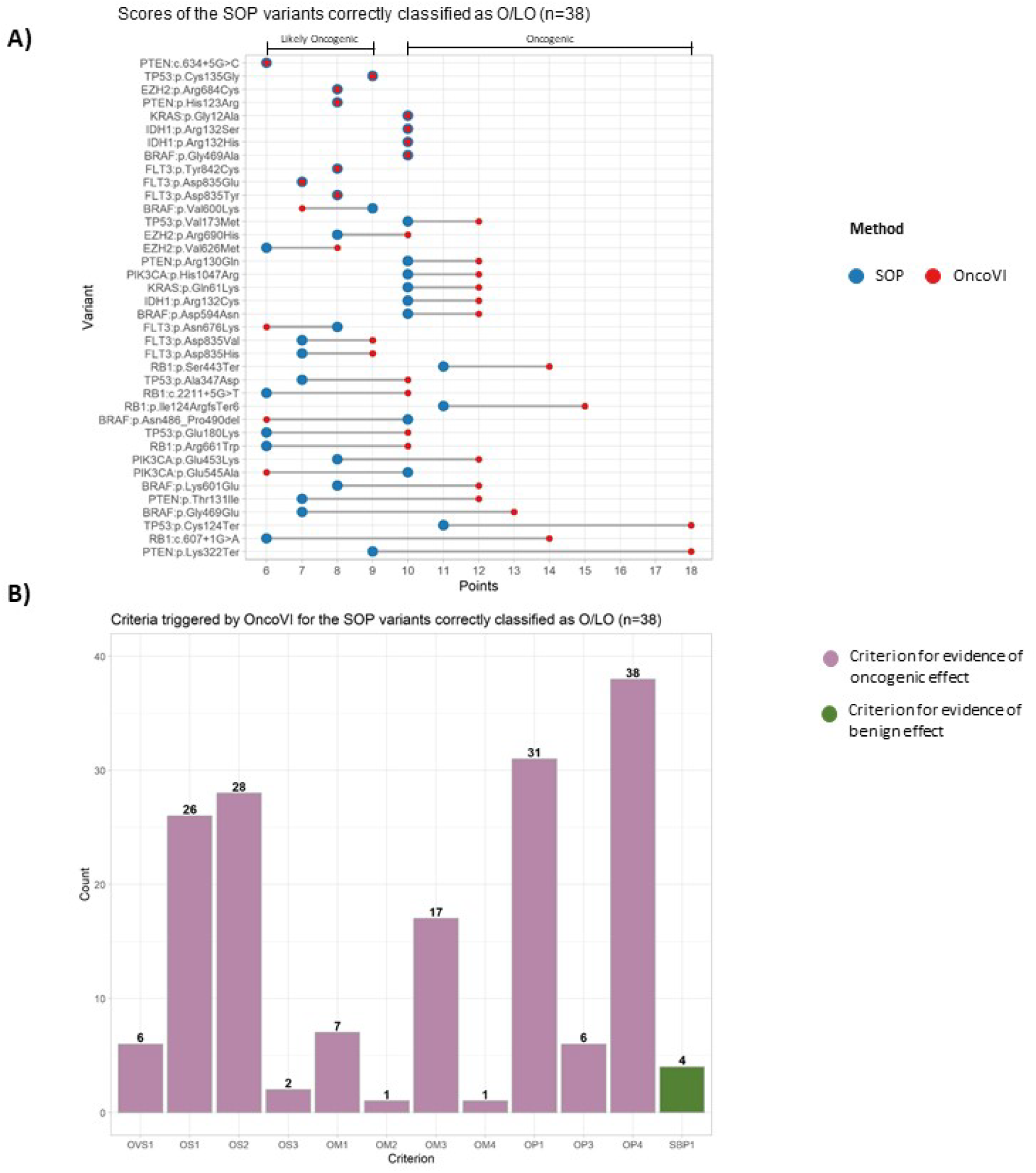
Results on the variants of the SOP data set correctly classified as O/LO. **A)** Dumbell plot of the 38 variants correctly classified as Oncogenic/Likely Oncogenic (O/LO) Horizontal bars indicate the label of the variants according to the SOP oncogenicity classification (i.e., scores≥10:Oncogenic, 10>score≥6:Likely Oncogenic). **B)** Barplot of the criteria triggered by OncoVI in the 38 variants correctly classified as O/LO. Criteria are sorted according to decreasing corresponding points. OVS1: Oncogenic Very Strong-1 (8 points), OS1: Oncogenic Strong-1 (4 points), OS2: Oncogenic Strong-2 (4 points), OS3: Oncogenic Strong-3 (4 points), OM1: Oncogenic Moderate-1 (2 points), OM2: Oncogenic Moderate-2 (2 points). OM3: Oncogenic Moderate-3 (2 points). OM4: Oncogenic Moderate-4 (2 points), OP1: Oncogenic Supporting-1 (1 point), OP3: Oncogenic Supporting-3 (1 point), OP4: Oncogenic Supporting-4 (1 point), SBP1: Somatic Benign Supporting-1 (−1 point).

**Supplementary Figure 2.**
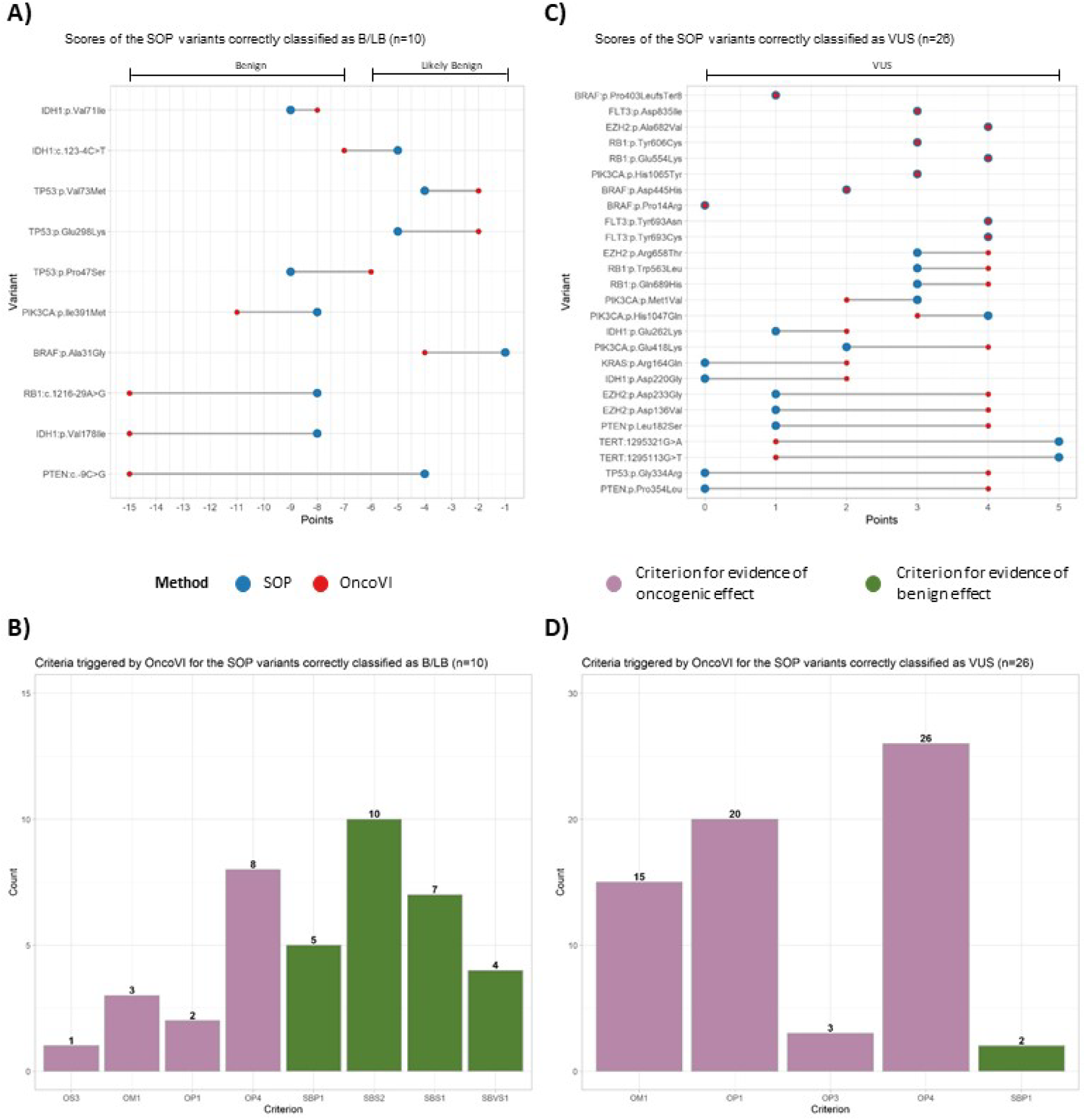
Results on the variants of the SOP data set correctly classified as B/LB and VUS. **A)** Dumbell plot of the 11 variants correctly classified as Benign/Likely Benign (B/LB) Horizontal bars indicate the label of the variants according to the SOP oncogenicity classification (O>score≥-6:Likely Benign, scores≤-7:Benign). **B)** Barplot of the criteria triggered by OncoVI in the ten variants correctly classified as B/LB, sorted according to decreasing corresponding points. **C)** Dumbell plot of the 26 variants correctly classified as variant of uncertain significance (VUS). Horizontal bar indicates the label of the variants according to the SOP oncogenicity classification (5>score≥0:VUS). **D)** Barplot of the criteria triggered by OncoVI in the 26 variants correctly classified as VUS. Criteria are sorted according to decreasing corresponding points. OS3: Oncogenic Strong-3 (4 points), OM1: Oncogenic Moderate-1 (2 points), OP1: Oncogenic Supporting-1 (1 point), OP3: Oncogenic Supporting-3 (1 point), OP4: Oncogenic Supporting-4 (1 point), SBP1: Somatic Benign Supporting-1 (−1 point), SBS2: Somatic Benign Strong-2 (−4 points), SBS1: Somatic Benign Strong-1 (−4 points), SBVS1: Somatic Benign Very Strong-1 (−8 points).

**Supplementary Figure 3.**
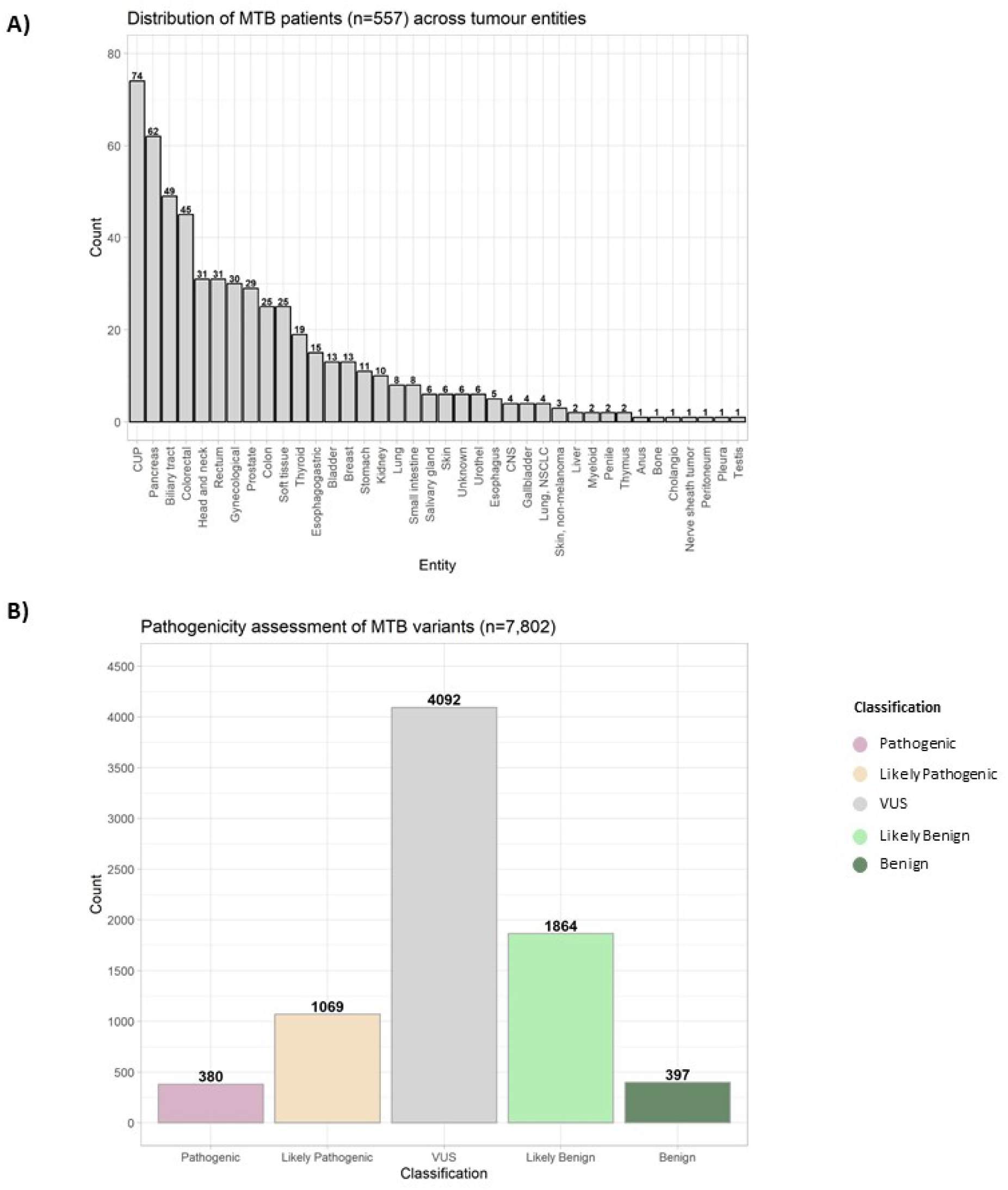
Molecular Tumour Board (MTB) data set of real-world routine diagnostic somatic variants. **A)** Distribution of the tumour entities associated with the 557 patients of the MTB cohort. CUP: cancer of unknown primary. CNS: central nervous system. Lung. NSCLC: Non-small-cell lung cancer. **B)** Distribution of the MTB pathogenicity assessment for all the 7,802 variants of the MTB data set. VUS: Variant of uncertain significance.

**Supplementary Figure 4.**
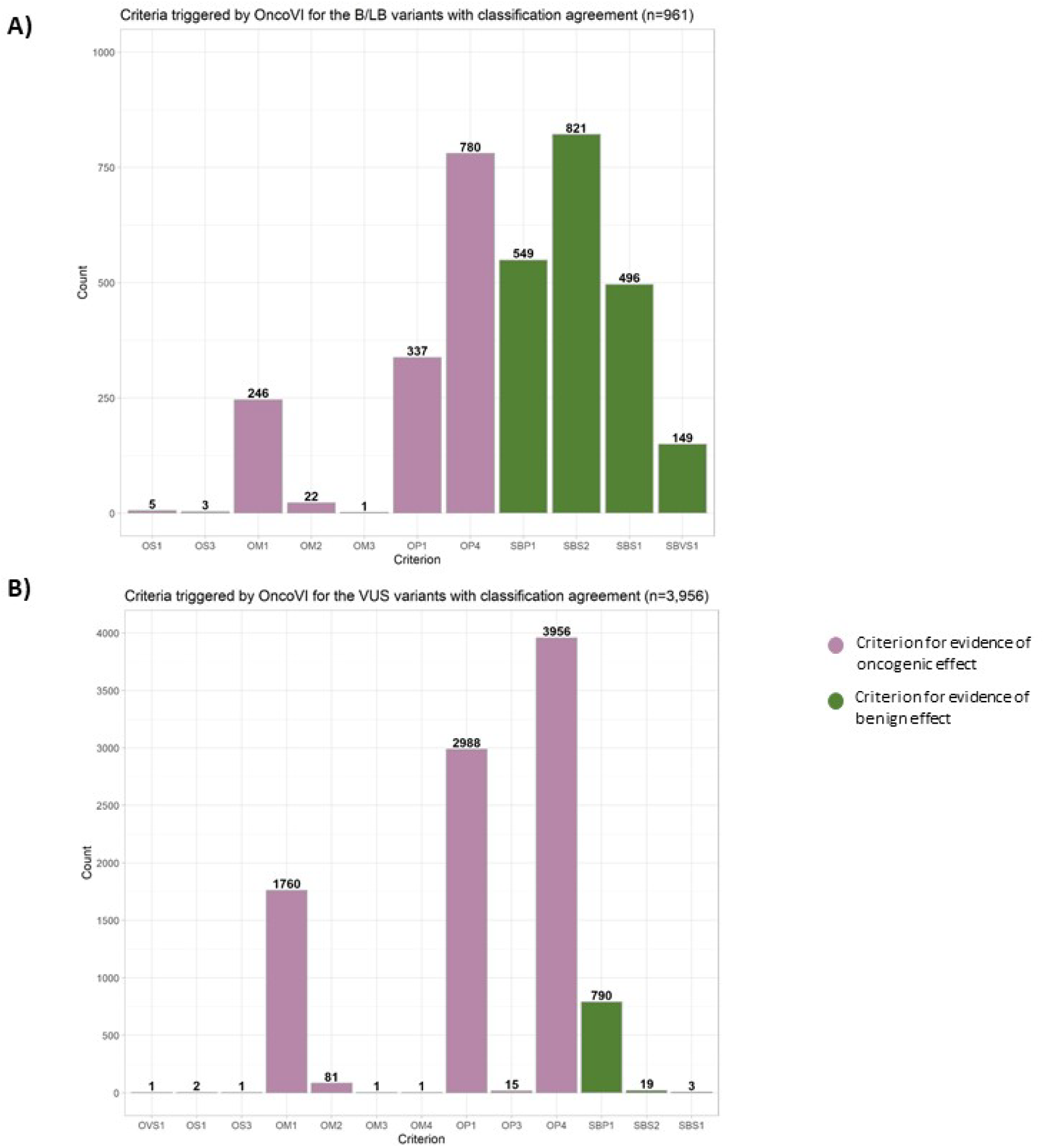
Criteria of the B/LB and VUS variants of the MTB data set with classification agreement. **A)** Barplot of the criteria triggered by OncoVI in the 961 variants classified as Benign/Likely Benign (B/LB) by both MTB and OncoVI Criteria are sorted according to decreasing corresponding points. **B)** Barplot of the criteria triggered by OncoVI in the 3,956 variants classified as variant of uncertain significance (VUS) by both MTB and OncoVI. Criteria are sorted according to decreasing corresponding points. OVS1: Oncogenic Very Strong-1 (8 points), OS1: Oncogenic Strong-1 (4 points), OS3: Oncogenic Strong-3 (4 points), OM1: Oncogenic Moderate-1 (2 points), OM2: Oncogenic Moderate-2 (2 points), OM3: Oncogenic Moderate-3 (2 points), OM4: Oncogenic Moderate-1 (2 points), OP1: Oncogenic Supporting-1 (1 point), OP3: Oncogenic Supporting-3 (1 point), OP4: Oncogenic Supporting-4 (1 point), SBP1: Somatic Benign Supporting-1 (−1 point), SBS2: Somatic Benign Strong-2 (−4 points), SBS1: Somatic Benign Strong-1 (−4 points), SBVS1: Somatic Benign Very Strong-1 (−8 points).

**Supplementary Figure 5.**
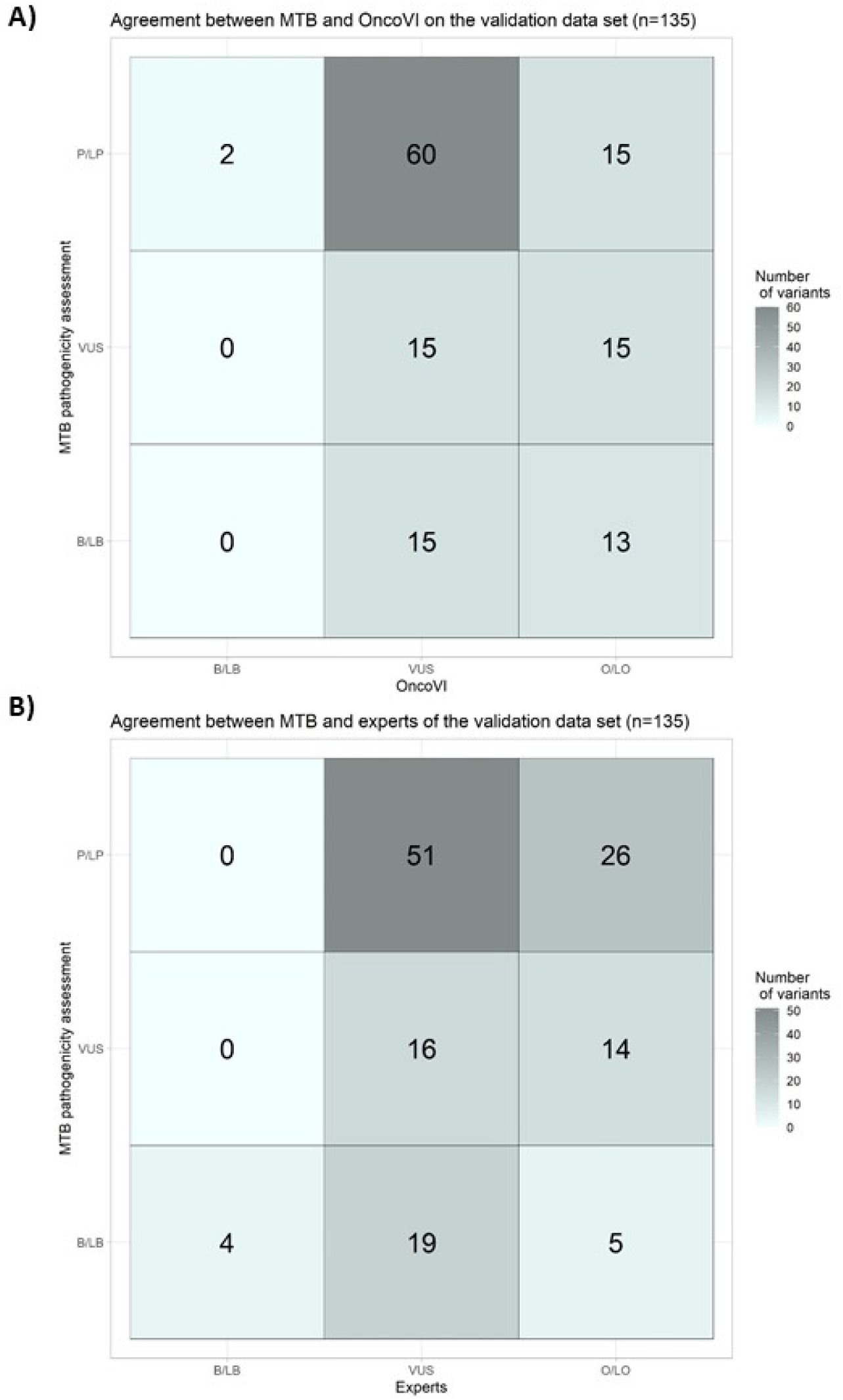
MTB variants of the validation data set. **A)** Confusion matrix of the agreement between OncoVl’s oncogenicity and the MTB pathogenicity in classifying the 135 somatic variants of the validation data set. B/LB Benign/Likely Benign. VUS: Variant of uncertain significance. O/LO: Oncogenic/Likely Oncogenic. PILP: Pathogenic/Likely Pathogenic. **B)** Confusion matrix of the agreement between MTB pathogenicity and expert re-assessment, based on the oncogenicity guidelines, in classifying, the 135 somatic variants of the validation data set.

**Supplementary Figure 6.**
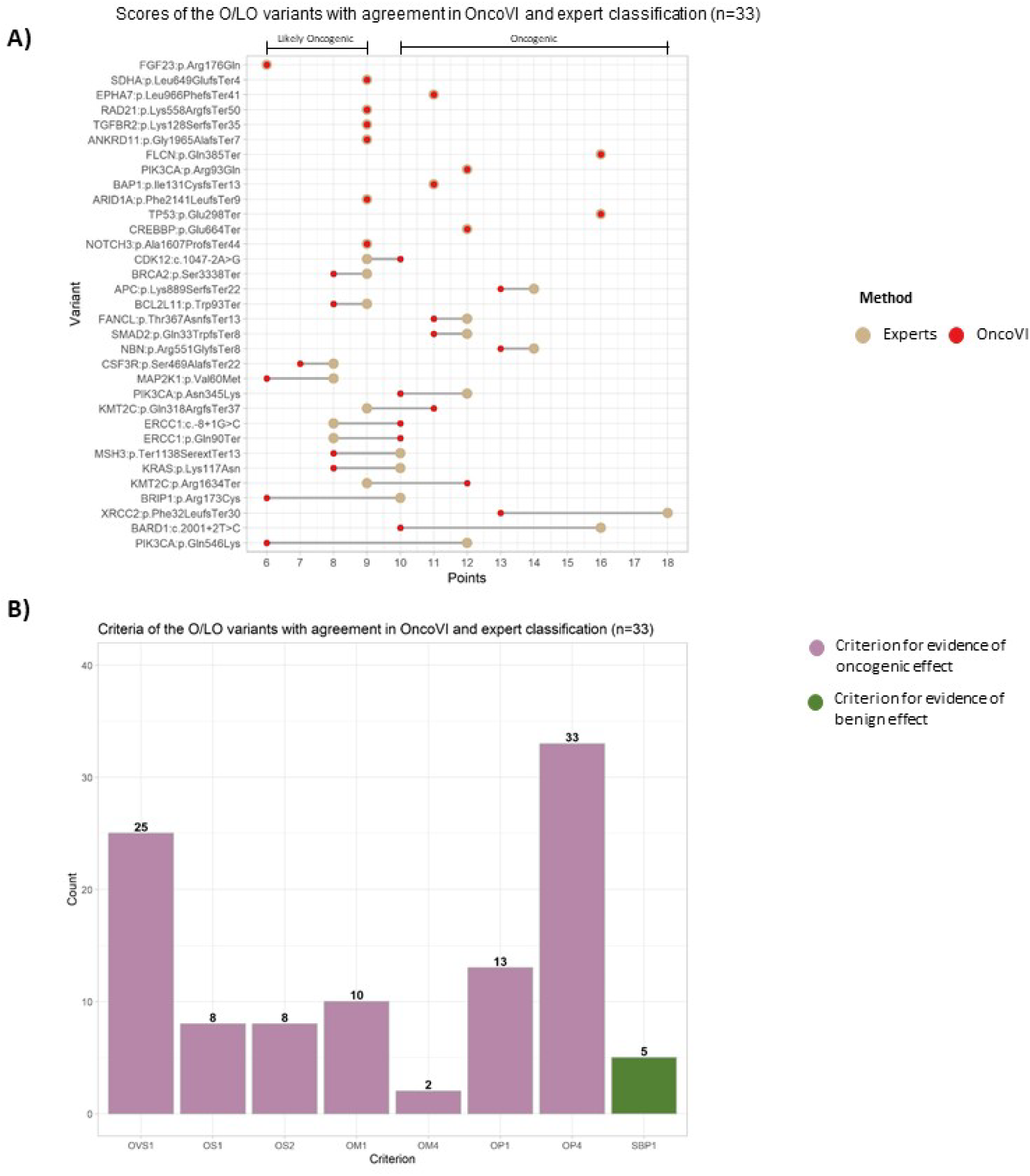
Results on the variants re-assessed as O/LO with agreement between expert and OncoVI classification. **A)** Dumbell plot of the 33 variants classified as Oncogenic/Likely Oncogenic (O/LO) by both experts and OncoVI. Horizontal bars indicate the label of the variants according to the expert classification (i.e.. scored ≥10:Oncogenic, 10>score≥6:Likely Oncogenic). **B)** Barplot of the criteria triggered by OncoVI in the 33 variants classified as O/LO by both experts and OncoVI. Criteria are sorted according to decreasing corresponding points. OVS1: Oncogenic Very Strong-1 (8 points), OS1: Oncogenic Strong-1 (4 points), OS2: Oncogenic Strong-2 (4 points), OM1: Oncogenic Moderate-1 (2 points), OM4: Oncogenic Moderate-4 (2 points), OP1: Oncogenic Supporting-1 (1 point), OP4: Oncogenic Supporting-4(1 point). SBP1: SomaticBenign Supporting-1 (−1 point).

**Supplementary Figure 7.**
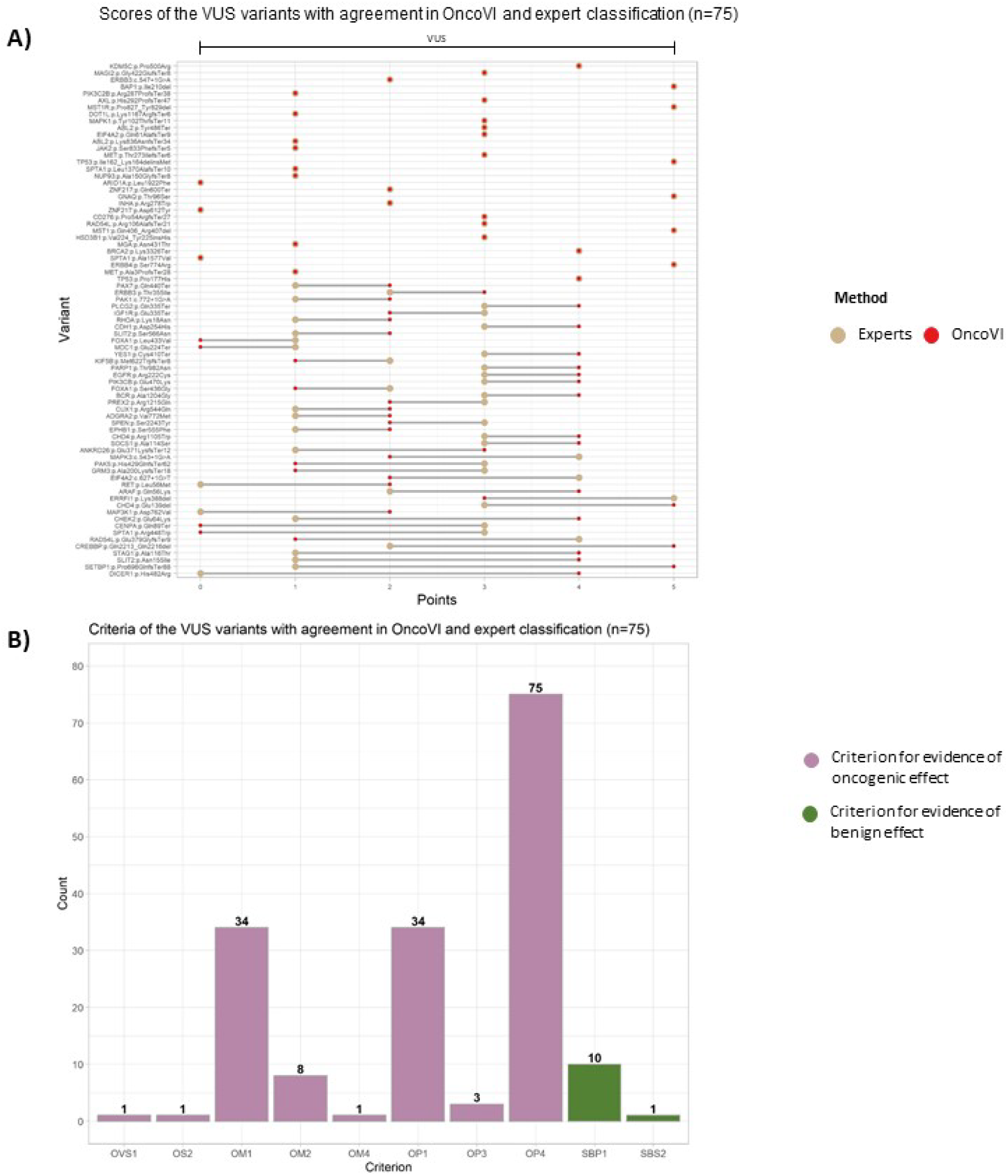
Results on the variants re-assessed as VUS with agreement between expert and OncoVI classification. **A)** Dumbell plot of the 75 variants classified as variant of uncertain significance (VUS) by both experts and OncoVI. Horizontal bar indicates the label of the variants according to the expert classification (5>score≥0:VUS) **B)** Barplot of the criteria triggered by OncoVI in the 75 variants classified as VUS by both experts and OncoVI. Criteria are sorted according to decreasing corresponding points OVS1: Oncogenic Very Strong-1 (8 points), OS2: Oncogenic Strong-2 (4 points). OM1: Oncogenic Moderate-1 (2 points). OM2: Oncogenic Moderate-2 (2 points), OM4: Oncogenic Moderate-4 (2 points), OP1: Oncogenic Supporting-1 (1 point), OP3: Oncogenic Supporting-3 (1 point), OP4: Oncogenic Supporting-4 (1 point), SBP1: Somatic Benign Supporting-1 (−1 point), SBS2: Somatic Benign Strong-2 (−4 points).

**Supplementary Figure 8.**
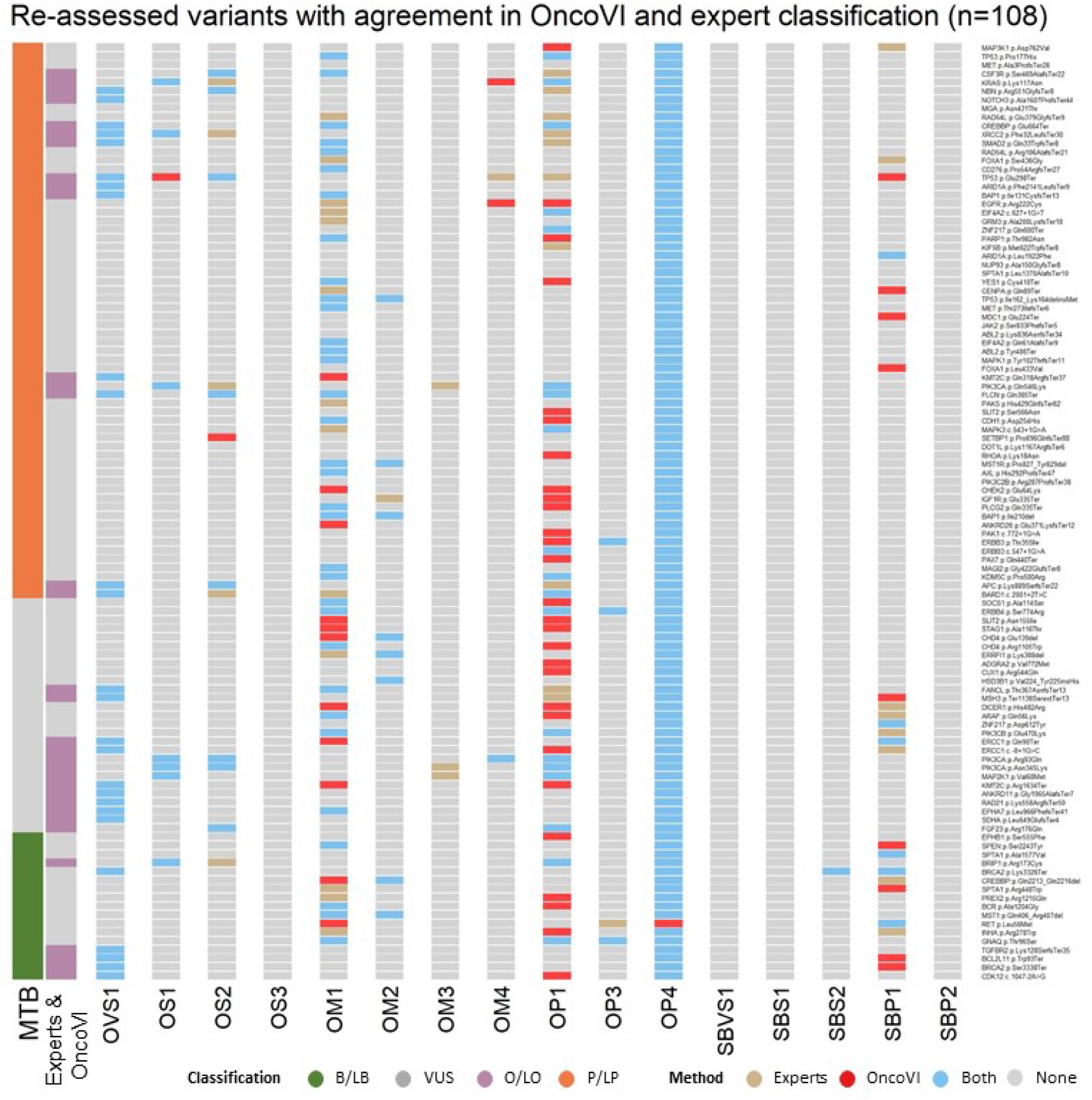
Re-assessed MTB variants with agreement between expert and OncoVI classification. Each row corresponds to one of the 108 variants with agreement between expert and OncoVI classification, each column corresponds to an assessed criterion. The colour of the cell indicates whether a criterion was triggered by the experts only (dark gold), OncoVI only (red), both (light blue) or none of the two (grey). The barplot on the left indicates the classification of each variant according to the Molecular Tumour Board (MTB), OncoVI, and expert assessment B/LB: Benign/Likely Benign, O/LO: Oncogenic/Likely Oncogenic, VUS: Variant of uncertain significance, P/LP: Pathogenic/Likely Pathogenic OVS1: Oncogenic Very Strong-1 (8 points), OS1: Oncogenic Strong-1 (4 points), OS2: Oncogenic Strong-2 (4 points), OS3: Oncogenic Strong-3 (4 points), OM1: Oncogenic Moderate-1 (2 points), OM2: Oncogenic Moderate-2 (2 points), OM3: Oncogenic Moderate-3 (2 points), OM4: Oncogenic Moderate-4 (2 points), OP1: Oncogenic Supporting-1 (1 point), OP3: Oncogenic Supporting-3 (1 point), OP4: Oncogenic Supporting-4 (1 point), SBVS1: Somatic Benign Very Strong-1 (−8 points), SBS1: Somatic Benign Strong-1 (−4 points), SBS2: Somatic Benign Strong-2 (−4 points), SBP1: Somatic Benign Supporting-1 (−1 point), SBP2: Somatic Benign Supporting-2 (−1 point).

## Notes

### Funding Statement

This study was funded by Deutsche Forschungsgemeinschaft (German Research Foundation) grant TRR 305 [projects Z01 (FF) and Z02 (ArHa)]. We express gratitude to patients, clinicians, technicians and institutions who contributed to the Molecular Tumour Board of the CCC Erlangen-EMN (Germany)

